# Risk scores for predicting HIV incidence among adult heterosexual populations in sub-Saharan Africa: a systematic review and meta-analysis

**DOI:** 10.1101/2021.09.28.21264246

**Authors:** Katherine M. Jia, Hallie Eilerts, Olanrewaju Edun, Kevin Lam, Adam Howes, Matthew L. Thomas, Jeffrey W. Eaton

**Affiliations:** MRC Centre for Global Infectious Disease Analysis, School of Public Health, Imperial College London, London, United Kingdom; Department of Population Health, The London School of Hygiene and Tropical Medicine, London, United Kingdom; Department of Mathematics, Imperial College London, London, United Kingdom; Joint Centre for Excellence in Environmental Intelligence, University of Exeter & Met Office, Exeter, United Kingdom

**Author notes:** Corresponding author: Jeffrey W. Eaton, MRC Centre for Global Infectious Disease Analysis, School of Public Health, Imperial College London, St. Mary’s Hospital Campus, Norfolk Place, London W2 1PG, United Kingdom. E-mail addresses of authors: KMJ.

**Keywords:** risk scores, HIV incidence, sub-Saharan Africa, adolescent girls and young women, risk factors for HIV incidence

## Abstract

**Introduction:** Several HIV risk scores have been developed to identify individuals for prioritised HIV prevention in sub-Saharan Africa. We systematically reviewed HIV risk scores to: (i) identify factors that consistently predicted incident HIV infection, (ii) review inclusion of community-level HIV risk in predictive models, and (iii) examine predictive performance.

**Methods:** We searched nine databases from inception until February 15, 2021 for studies developing and/or validating HIV risk scores among the heterosexual adult population in sub-Saharan Africa. Studies not prospectively observing seroconversion or recruiting only key populations were excluded. Record screening, data extraction, and critical appraisal were conducted in duplicate. We used random-effects meta-analysis to summarise hazard ratios and the area under the receiver-operating characteristic curve (AUC-ROC).

**Results:** From 1563 initial search records, we identified 14 risk scores in 13 studies. Seven studies were among sexually active women using contraceptives enrolled in randomised-controlled trials, three among adolescent girls and young women (AGYW), and three among cohorts enrolling both men and women. Consistently identified HIV prognostic factors among women were younger age (pooled adjusted hazard ratio: 1.62 [95% Confidence Interval: 1.17, 2.23], compared to above-25), single/not cohabiting with primary partners (2.33 [1.73, 3.13]) and having sexually transmitted infections (STIs) at baseline (HSV-2: 1.67 [1.34, 2.09]; curable STIs: 1.45 [1.17; 1.79]). Among AGYW only STIs were consistently associated with higher incidence, but studies were limited (n=3). Community-level HIV prevalence or unsuppressed viral load strongly predicted incidence but were only considered in three of 11 multi-site studies. The AUC-ROC ranged from 0.56 to 0.79 on the model development sets. Only the VOICE score was externally validated by multiple studies, with pooled AUC-ROC 0.626 [0.588, 0.663] (*I*^*2*^: 64.02%).

**Conclusions:** Younger age, non-cohabiting, and recent STIs were consistently identified as predicting future HIV infection. Both community HIV burden and individual factors should be considered to quantify HIV risk. However, HIV risk scores had only low-to-moderate discriminatory ability and uncertain generalisability, limiting their programmatic utility.

Further evidence on the relative value of specific risk factors, studies populations not restricted to ‘at-risk’ individuals, and data outside South Africa will improve the evidence base for risk differentiation in HIV prevention programmes.

**PROSPERO Number:** CRD42021236367

## Introduction

Efficiently identifying populations and individuals at high risk of HIV infection and linking them to effective HIV prevention is essential for continued progress towards ending HIV as a public health threat [1]. Differentiating HIV prevention based on risk of infection is especially important for interventions that are expensive and intensive for both the client and the health system, such as daily oral pre-exposure prophylaxis (PrEP) [2-5]. Identifying those at highest risk for infection is most difficult in sub-Saharan Africa, where 58% of the 1.5 million global new infections in 2020 occurred [6], and a large proportion of new infections were through heterosexual transmission among the general population [7].

Several HIV incidence risk scores have been proposed as prognostic tools for identifying individuals at high risk for HIV infection in sub-Saharan Africa [8, 9]. HIV risk scores combine data on multiple prognostic factors into a single score that summarises an individual’s risk for infection. Certain interventions might be offered, or restrict eligibility to, individuals with scores above a specified threshold [10]. An optimal threshold maximises the share of incident infections among the higher risk group while minimizing the total proportion classified as such, but there is typically a trade-off between these. Risk scores are empirically derived using data from large-scale, longitudinal studies like HIV randomised controlled trials (RCTs) and cohort studies that collect comprehensive HIV prognostic factors spanning the behavioural, sociodemographic, partnership domains among HIV negative adults and prospectively measure HIV incidence, usually within 1 year or less after the baseline risk assessment. Generalisability is validated by applying the risk score to independently collected data and studying how well the score discriminates those who subsequently acquire HIV.

Recently, national HIV programmes have focused on prioritising interventions to geographic areas with high HIV burden [1], but not widely implemented risk scoring tools to differentiate individual-level access to HIV interventions. The geographically focused strategy is epidemiologically justified for two reasons: high HIV burden indicates previous high HIV risk, and, secondly, high HIV prevalence or unsuppressed HIV viraemia implies greater exposure to HIV infection among those currently at risk [11, 12]. This community-level exposure does not fit naturally into the individual-level risk framework of risk scoring.

Mathematical modelling has demonstrated that considering both geographic location and risk populations in prioritising of HIV prevention improves the efficiency and cost-effectiveness relative to only one dimension [13]. The new Global AIDS Strategy 2021-2026 embraces this approach—recommending that HIV prevention is prioritised for various population groups differentiated according to thresholds for the local HIV incidence [14].

For example, for adolescent girls and young women (AGYW), the strategy recommends prioritisation of services to those at high risk based on: (i) the subnational annual incidence greater than 3%, or (ii) an incidence of 1-3% and self-reported high-risk behaviours or recent sexually transmitted infection (STI) [14].

We conducted a systematic review of HIV risk score tools in sub-Saharan Africa to explore this from both perspectives. Firstly, to motivate improved modelling of HIV incidence and prioritising of HIV prevention, we sought to identify prognostic factors from the HIV risk score literature that stratify population HIV risk, and the ability of these factors to discriminate HIV incidence within a population. Secondly, we queried the extent to which HIV risk scores considered community-level HIV prevalence or population viraemia as a predictor in prognostic models for individual HIV incidence risk. Specifically, we searched literature for studies that either developed or validated a HIV incidence risk score model among adult heterosexual populations, and analysed the data to: (i) identify risk factors that have consistently shown strong effects on HIV incidence across different models and settings, (ii) evaluate whether community-level HIV prevalence has been considered as a determinant of HIV risk in risk score development, and (iii) examine the efficiency of risk scores in differentiating high- and low-risk individuals quantified by the area under the receiver-operating characteristic curve (AUC-ROC).

## Methods

### Search strategy

We searched for studies that developed and/or validated the HIV incidence risk scores among adult heterosexual populations of sub-Saharan Africa. Specific inclusion criteria were: (i) development and/or validation of any predictive multivariable model (“risk score”) with prospectively measured HIV incidence as the main outcome (i.e., documented HIV-negative status at baseline), (ii) enrolled from adult heterosexual populations and (iii) conducted in sub-Saharan African countries. Studies were excluded if: (i) HIV seroconversions were not determined by a HIV negative test result at baseline followed by a positive or negative result during follow-up, (ii) study populations were key or selected populations only (men who have sex with men, female sex workers, pregnant women, serodiscordant couples, HIV-exposed infants, people who inject drugs).

Keywords, synonyms, and related terms covered the domains of “sub-Saharan Africa”, “HIV/AIDS”, and “risk score”. The full electronic search strings for all databases are available in the Supplementary Material (Appendix I). No restrictions were imposed on the types nor years of publications; however, only publications written in English were included.

### Sources of information

Nine databases were searched: MEDLINE, Embase, Global Health, PsycINFO, Maternity & Infant Care Database, CINAHL (EBSCO), Scopus, Cochrane Library, the Web of Science, on 15^th^ February 2021.

### Study selection

Titles and abstracts were independently screened by two reviewers for eligibility against the inclusion and exclusion criteria. Discrepancies were resolved by either consensus after discussion or decision of a third reviewer. After abstract screening, full texts were reviewed for inclusion by two independent reviewers. Reasons were provided for any exclusion of studies at this stage. Again, any discrepancies in decisions or reasons were resolved through discussion or by a third reviewer. Abstract screening, full text review, and data extraction were conducted by KMJ, HE, OE, KL, AH, and MJT.

### Data extraction and risk of bias assessment

Data were extracted by two independent reviewers, with discrepancies resolved through discussion. We referred to the Critical Appraisal and Data Extraction for Systematic Reviews of Prediction Modelling Studies (CHARMS) Checklist when creating the data extraction form (Appendix II).^[15]^ After extraction, two reviewers assessed the risks of bias for each study independently using the Prediction Model Risk of Bias (PROBAST) assessment tool checklist,[16] under the four domains ‘Population’, ‘Predictor’, ‘Outcome’ and ‘Analysis.’ A domain where one or more criteria was/were not fulfilled might be judged as “high risk of bias,” whereas a study with one (or more) domain(s) at “high risk of bias” would be judged as having an overall “high” risk of bias.

### Data synthesis and reporting

We aimed to identify significant and measurable prognostic factors that define high risk groups or individuals for prioritised HIV prevention. We first summarised the key characteristics, setting, and study population(s) of each included study, and whether it developed a risk score, externally validated a score, or both. A development study could conduct internal validation by using re-sampling methods (bootstrap or cross-validation) to estimate the AUC-ROC, or by splitting the sample into training and testing sets; external validation where the risk score was applied to a different study population than which it was originally derived can be performed in the same analysis or by others in follow-up studies. We then assessed the importance of each predictor by examining (i) the number of times it was included in the final risk prediction model of a model development study, (ii) the summary of the adjusted and unadjusted effect size estimates. Finally, we summarised the area under the receiver operating characteristic curve (AUC-ROC), proportion identified “high risk” by each score, and the corresponding HIV incidence in the high-risk group, to assess the risk scores discrimination and compared them across settings to examine generalisability.

Overall summary effect size estimates (and the 95% confidence interval) for predictors were estimated by a random effects model. Estimates were pooled for both the adjusted and unadjusted effects because adjusted effects were only available in studies that included the particular predictors in the multivariable models (due to significant univariate association), risking biasing summary estimates away from null. Between-study variance were reported with the I^2^ statistics to evaluate the heterogeneity. Random effects meta-analysis based on the inverse variance method with Sidik-Jonkman estimator for between-study variance was done in R (version 4.0.3) [17] using the packages “meta” and “metafor” [18, 19]. Meta-analysis for AUC-ROC was performed using methods described by Zhou and colleagues in Medcalc (version 19.8) [20, 21]. Forests plots and funnel plots were created using the package “meta” and Medcalc respectively.

The systematic review protocol was pre-registered on PROSPERO (CRD42021236367) [22]. We referred to the PRISMA (Preferred Reporting Items for Systematic Reviews and Meta-Analyses) checklist for presenting the review [23].

## Results

Database searches identified 2029 records; 466 duplicates were removed and 1563 titles and/or abstracts were screened, of which 25 studies were retained for full-text screening. One additional conference abstract was available after initial screening, resulting in 13 studies (9 peer-reviewed articles, 2 posters, 1 editorial letter, 1 abstract) that met the inclusion criteria and were included in this review (Figure 1) [8, 9, 24-34]. Critical appraisal according to the PROBAST checklist concluded that one out of twelve models developed and two out of nine validated were of low risk of bias (Figure S1, Table S3 & S4).

**Figure 1.**
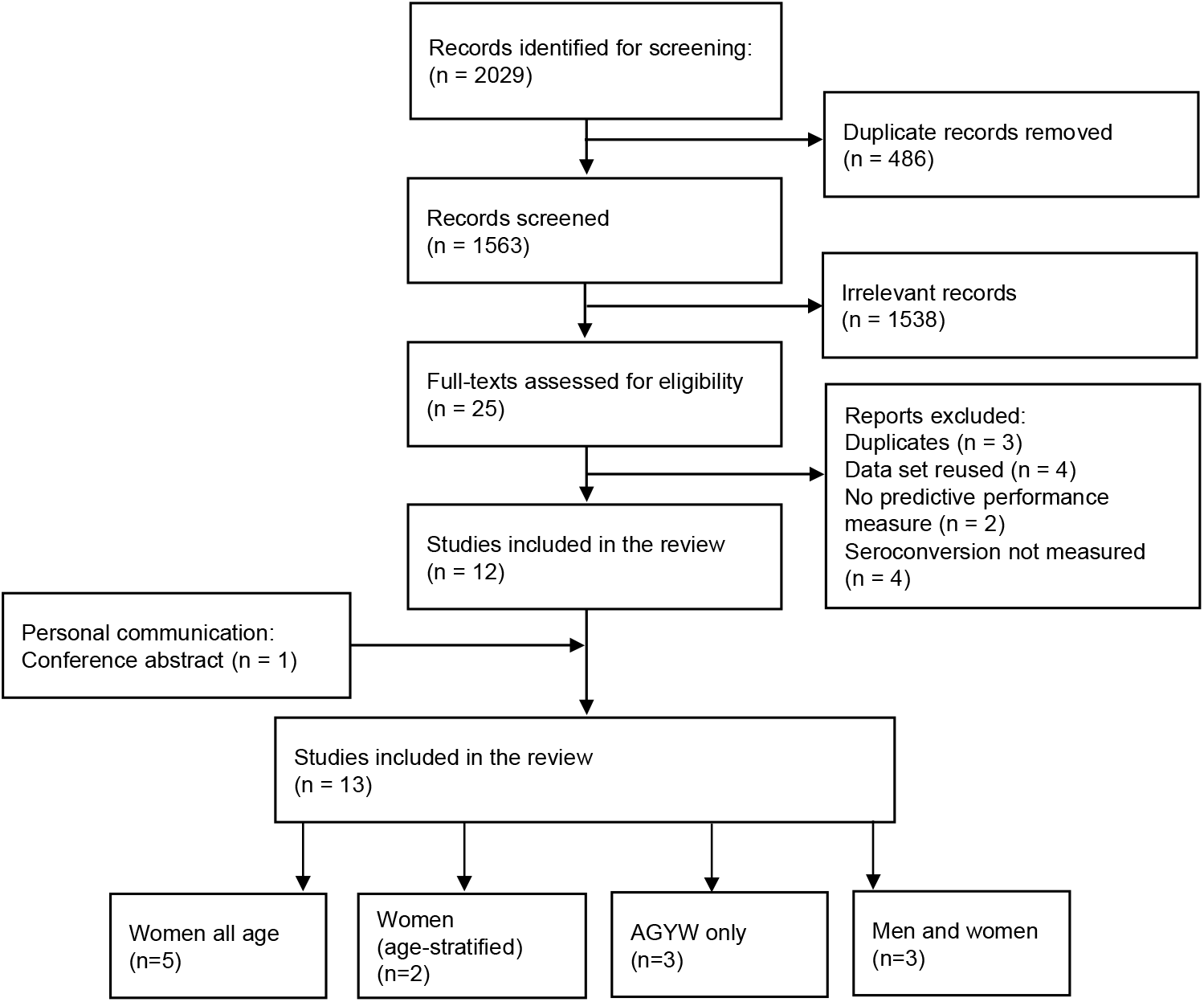
HIV risk score study selection. **n:** number; **AGYW:** adolescent girls and young women (AGYW)

Inadequate adjustment for over-fitting or model optimism was common among the development studies (6 out of 9). For the validation studies, inadequate sample size (4 of 9) and missing predictors (5 of 9) were common limitations. Among studies that reported information about loss-to-follow-up and incomplete data (9 of 15 studies; Table S5), the proportion of enrolled participants included in final analysis ranged from 80% to over 95% in the RCTs (except for one) and 60% in RCCS open-cohort study. The three studies with data from population cohorts used imputation to account for missing data and Ayton imputed unavailable predictor variables (Table S5).

## Study populations

Studies were conducted in South Africa (n=10), Uganda (n=4), Malawi, Zimbabwe (n=3), Kenya (n=2), Zambia (n=1), and Tanzania (n=1). Three enrolled multi-country study populations, and eight were multi-site within one country (Table 1). A total of 134,423 individuals (301,820 person-years) were included in the studies, among whom 28.0% (N=37599; 73,955 person-years) were from South Africa. One study in Uganda and Kenya accounted for 56% of all participants (75,558 individuals) [33]. The mean HIV incidence was 4.82 per 100 person-years in studies conducted in South Africa and 2.34 per 100 person-years elsewhere. Incidence and risk factor data were collected before 2012 for seven studies (mean incidence: 4.61 per 100 person-years) and after 2012 for six (mean incidence: 3.16 per 100 person-years). The majority (10 of 13) were among women only, of which three were restricted to young women under age 25 or lower; 3 included women and men aged 15–49 years or 15 years and older. The majority (10 of 13) were RCTs or quasi-experimental studies that restricted recruitment and/or eligibility to specific at-risk population groups: (i) sexually active, contraception-seeking women who attended the family-planning, STIs or research clinics (7 of 10, all RCTs) [8, 9, 24-28], or (ii) school-attending AGYW (3 of 10) [29-31]. The remaining three were large-scale cohort studies or community trials that recruited all consenting members within the communities [32-34].

**Table 1.**
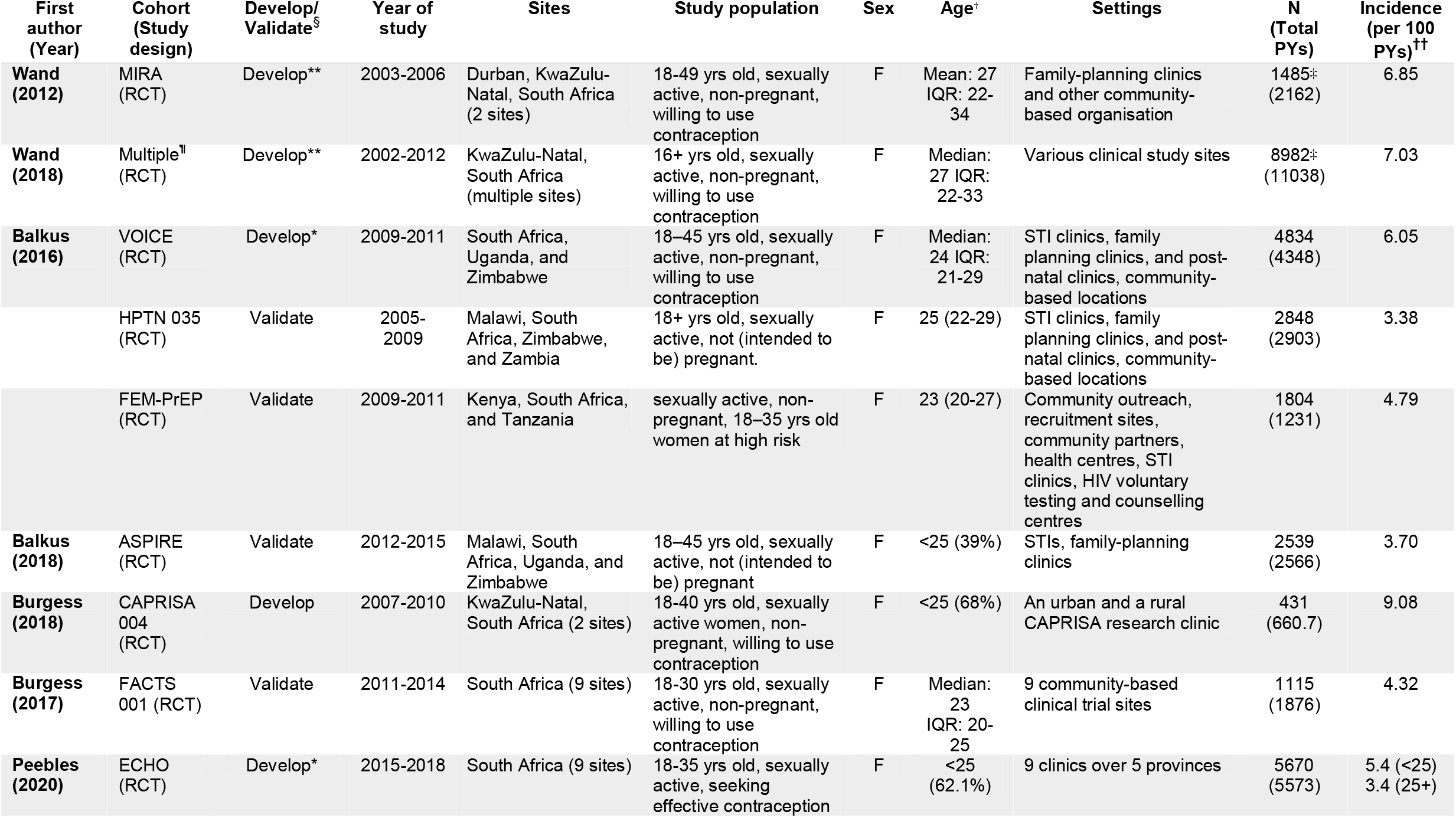
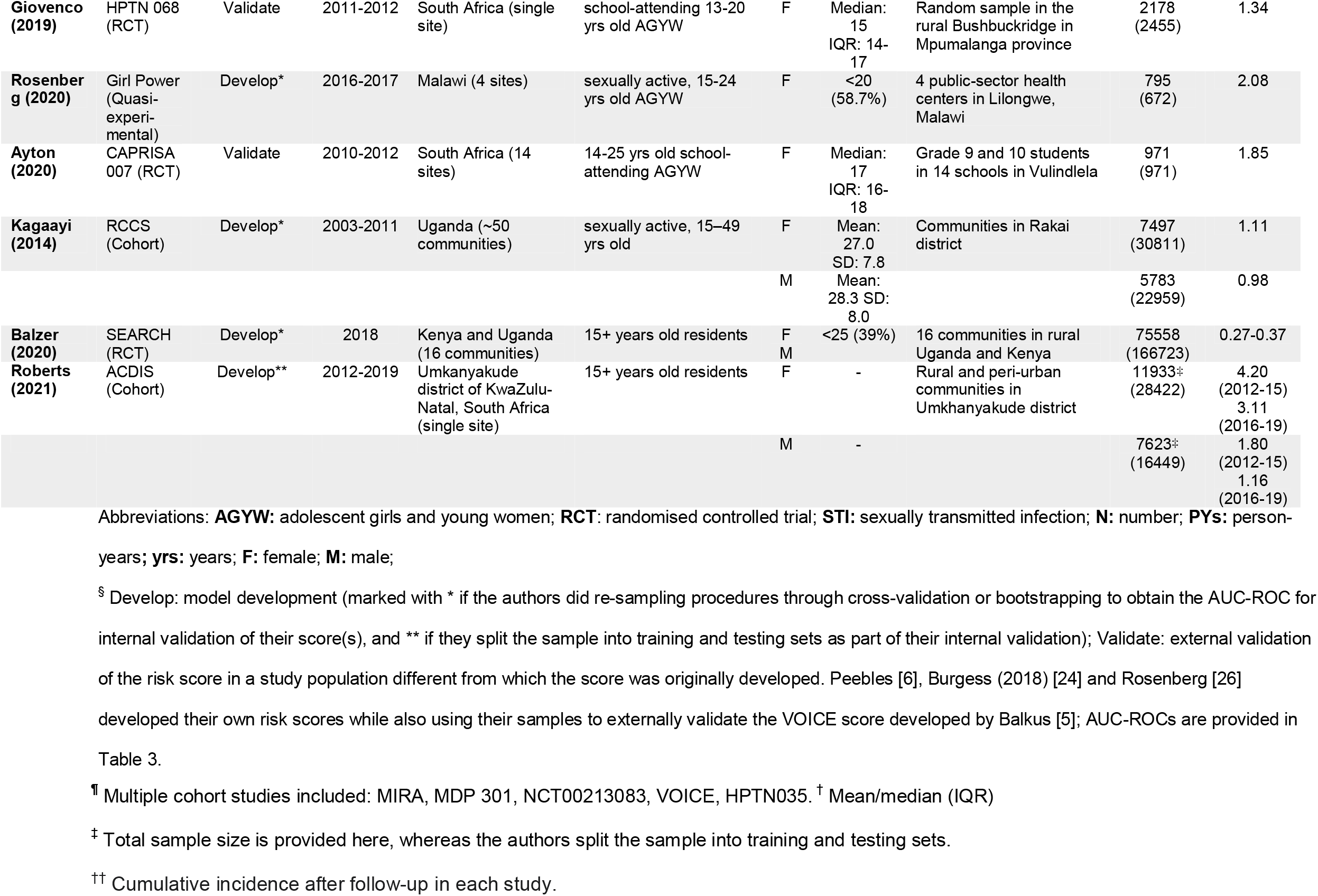
Characteristics of the cohorts from which HIV risk scores were developed and/or validated.

Geographic locations, study periods, age groups, and settings are given in Table 1.

## Factors included in HIV risk scores

Nine studies reported on development of 14 HIV risk scores, involving screening and model selection for baseline predictors of HIV incidence (Table 1). Balzer used a machine learning approach, specifically the Super Learner ensemble model method [35], which did not yield effect estimates for individual risk factors [33]. Final regression results were also not available for Roberts (abstract only) [34]. For the remaining studies, Table 2 reports the predictors considered for inclusion and retained in the final model for each independently developed score. The ‘*retainment ratio*’ reports the number of times a risk factor was retained in the final score relative to the number of times it was considered as a ‘*candidate*’ predictor, tabulated separately for risk scores for women of all ages and for AGYW only study populations.

**Table 2.**
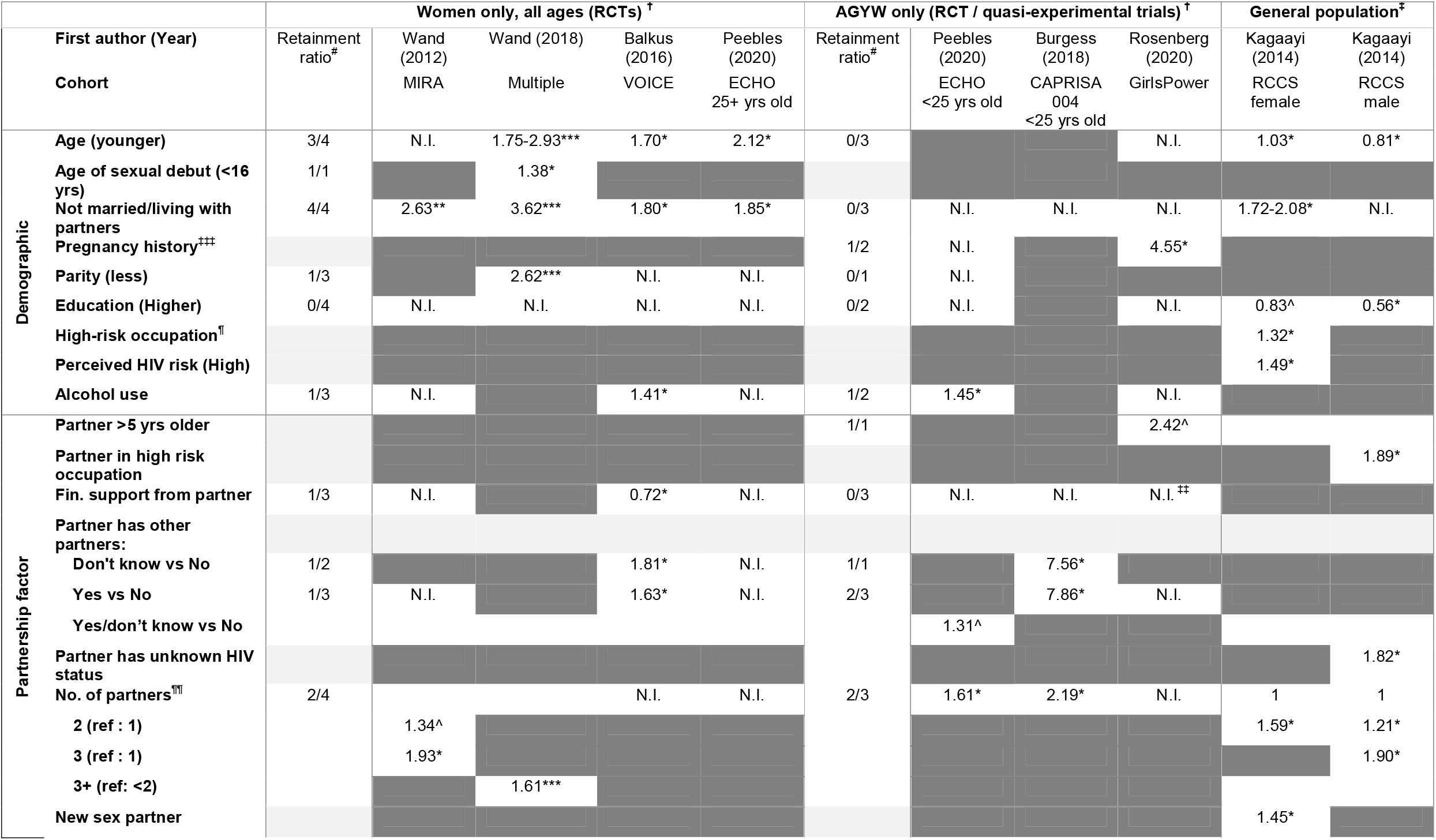
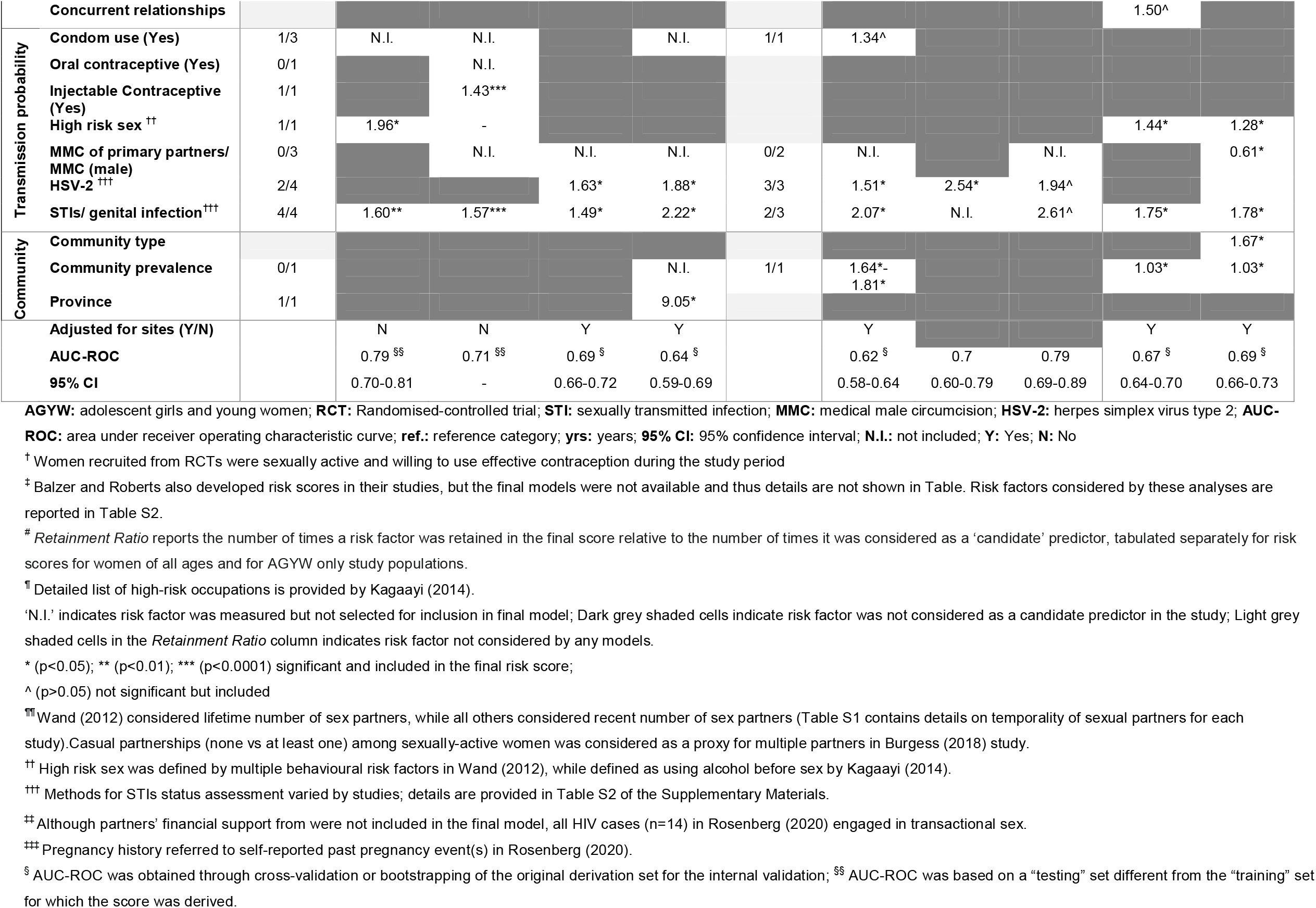
Risk factors retained in the final HIV risk score models and adjusted effect estimates.

All of the four risk scores for all age, sexually active, contraceptive-seeking women were developed using RCT data in South Africa (VOICE included data from other countries but 81% of study participants were from South Africa). Factors retained in all or three of four final models were: not being married or cohabiting with primary partner (pooled adjusted hazard-ratio [aHR] 2.33; 95% CI [1.73, 3.13]; Table S6); younger age (pooled aHR: 1.62 [1.17, 2.23]; less than 25 years old except for Peebles [9] at 27 years), and curable STIs at baseline (pooled aHR 1.45 [1.17, 1.79]) (Figure 2). Human Simplex Virus – 2 (HSV-2; pooled aHR 1.67 [1.34, 2.09]) and multiple sexual partners (Pooled aHR: 1.62 [1.27, 2.07]) were included in two of four risk scores. Other demographic, partnership, biological or community factors were either seldom considered as candidate predictors or only retained in one or fewer risk scores (Table 2). Among unselected candidate predictors, educational attainment, employment (or earning own income) and coital frequency were considered by all four studies but not retained in any of the final models (Table S1, Supplementary Materials).

**Figure 2.**
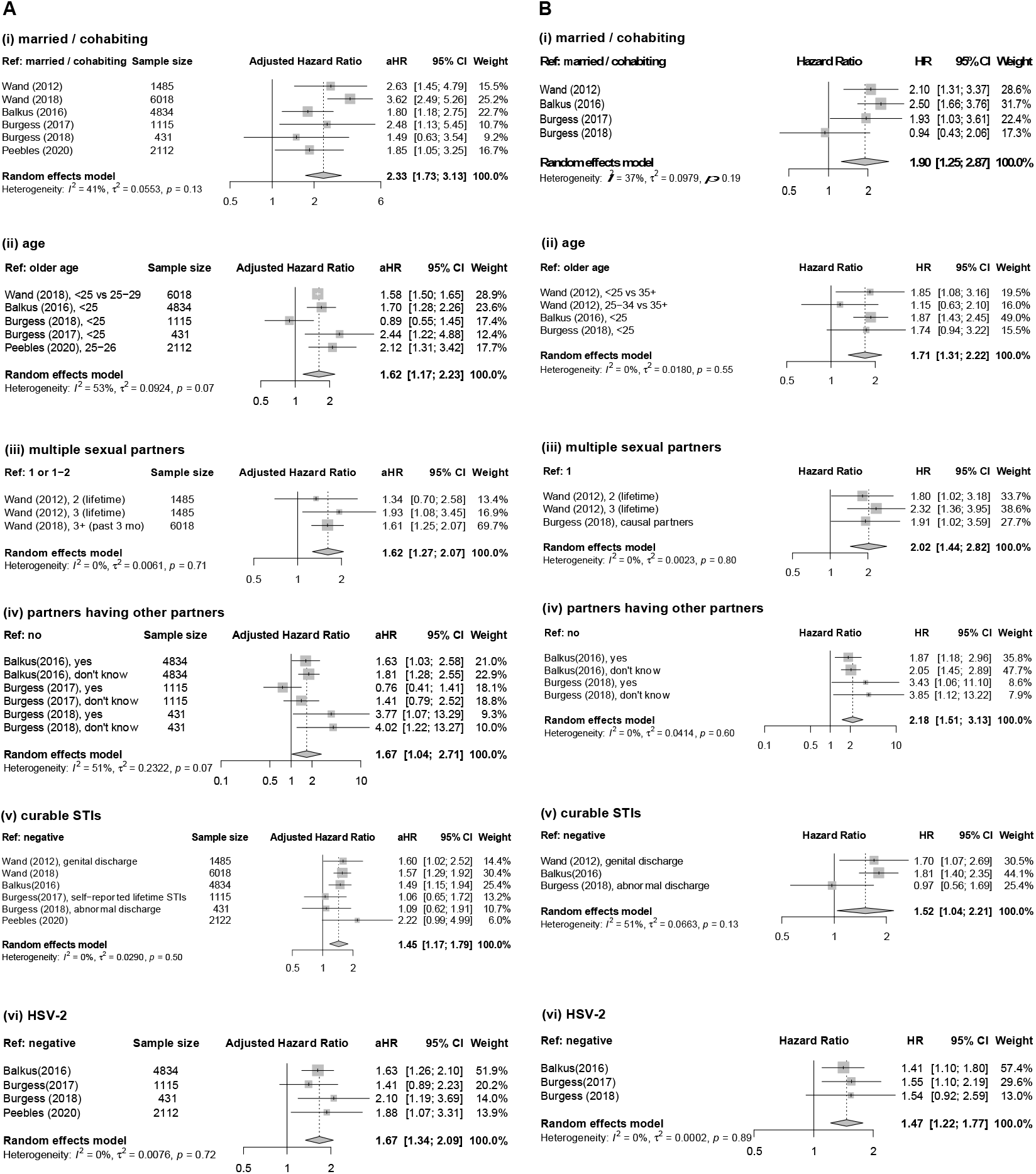
Forest plots of risk factor estimates among women in general. Adjusted (A) and unadjusted (B) effects were pooled together for: (i) marital / cohabiting status, (ii) age, (iii) number of sexual partners, (iv) partners having other partners, (v) curable sexually transmitted infection (STIs) and (vi) HSV-2. Abbreviations: **Ref:** reference category; **HR:** hazard ratio; **aHR:** adjusted hazard ratio; **95% CI:** 95% confidence interval; **STIs:** sexually transmitted infections; **HSV-2:** herpes simplex virus type 2.

Three risk scores were developed specifically for sexually active AGYW (aged 13-24, varying across studies) (Table 2). HSV-2 was the only factor selected in all three (pooled aHR: 1.77 [1.24; 2.54]). Factors selected in two of three models were curable STIs (pooled aHR: 2.14 [1.40; 3.25]), having multiple partners (pooled aHR: 1.76 [1.19; 2.60]) and partner having other sexual partners (pooled aHR: 2.35 [0.48; 11.53]) (Figure 3). Being not married/cohabitating was not selected in any final models, unlike the models for all age contraceptive-seeking women where it was selected by all models.

**Figure 3.**
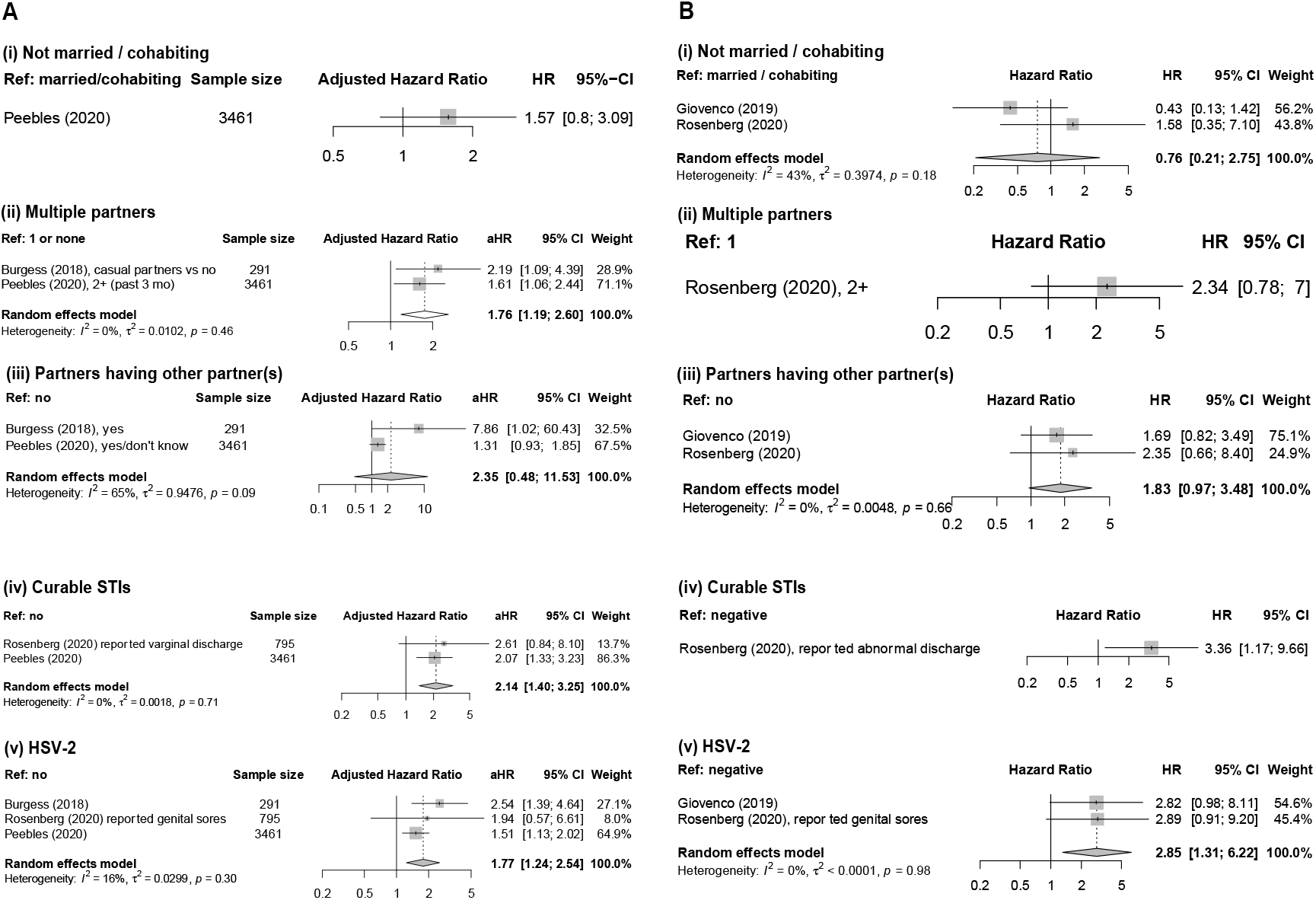
Forest plots of risk factor estimates among adolescent girls and young women (AGYW). Adjusted (A) and unadjusted (B) effects were pooled together for (i) marital / cohabiting status (ii) number of sexual partners, (iii) partners having other partners, (iv) curable sexually transmitted infection (STIs) and (v) HSV-2. Abbreviations: **Ref:** reference category; **HR:** hazard ratio; **aHR:** adjusted hazard ratio; **95% CI:** 95% confidence interval; **STIs**: sexually transmitted infections; **HSV-2:** herpes simplex virus type 2.

In summary, not being married or cohabiting was consistently identified and had the largest effect size estimates in studies among all-aged adult women. For AGYW presence of other STIs was most consistently selected. Of the remaining predictors, occupation, self-perceived HIV risk, partners’ occupation, having new partners, engaging in high-risk sex (e.g., under alcohol use), knowledge of partner’s HIV status showed significant associations but were seldom assessed [32].

## Inclusion of community HIV prevalence

Only three of 11 multi-site studies considered community-level HIV prevalence as a covariate and all cases where considered it was selected into one or more of the final models [9, 32, 34]. In Peebles [9], compared to residing in a community with 10-15% HIV prevalence, those in a community with 16-20% prevalence had an aHR of 1.64 [1.08, 2.48], 1.71 [0.99, 2.96] for 21-25% prevalence, and 1.81 [1.03, 3.19] for 26-30%. Similarly, in Kagaayi [32], an aHR of 1.03 was associated with each percentage-point increment in community prevalence for both male [0.99,1.07] and female [1.01, 1.06]. Roberts [34] also found community HIV prevalence and unsuppressed viral load to be highly predictive, but aHRs were not available.

## Predictive performance of the risk scores

We identified 14 risk scores from nine model development studies (three models developed by Balzer were considered separately) [33] (Table 3). Most studies used baseline predictors to predict incidence infections observed during the following one year, with some extending to 18 months or two years (Table 3).

**Table 3.**
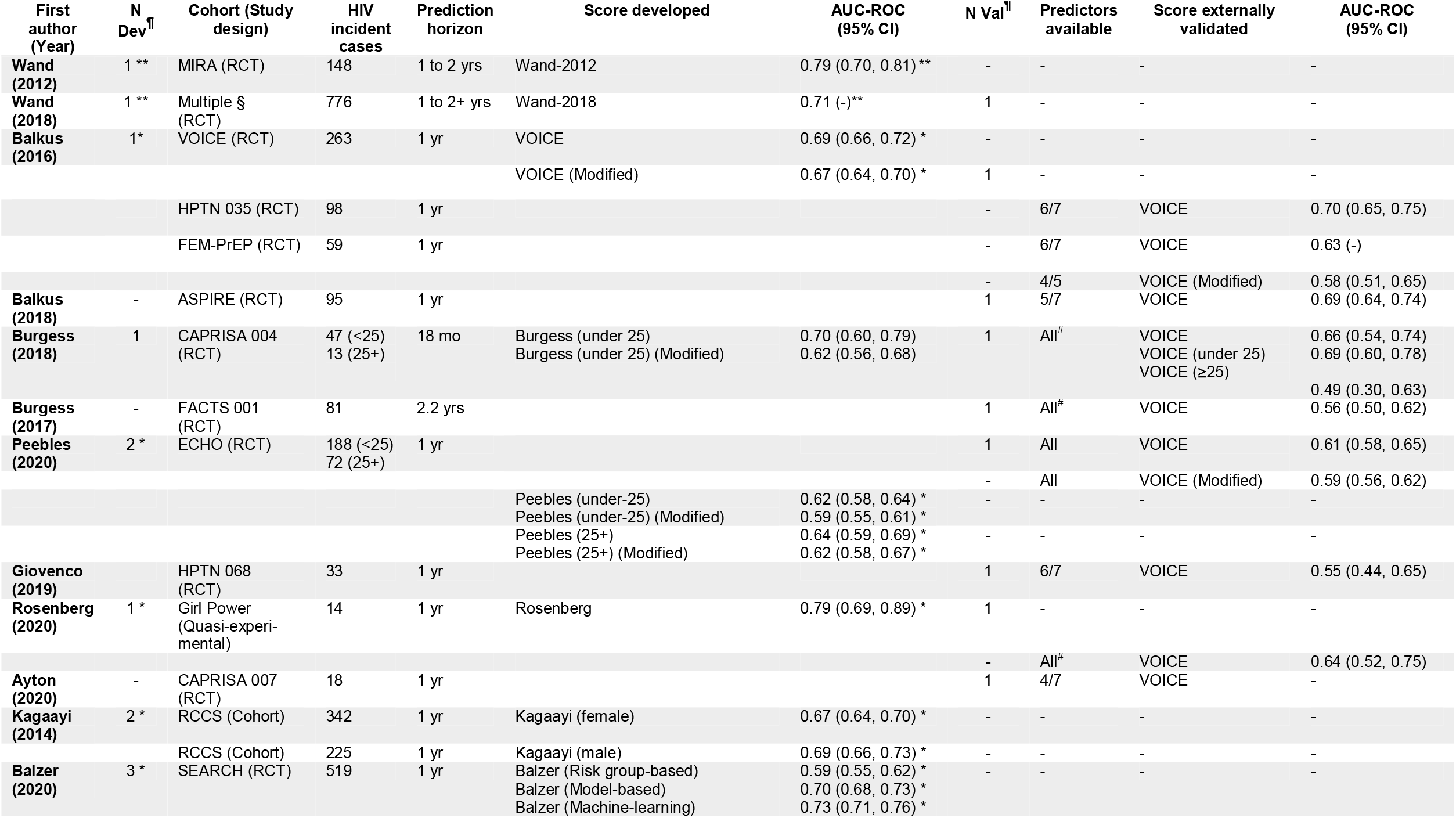
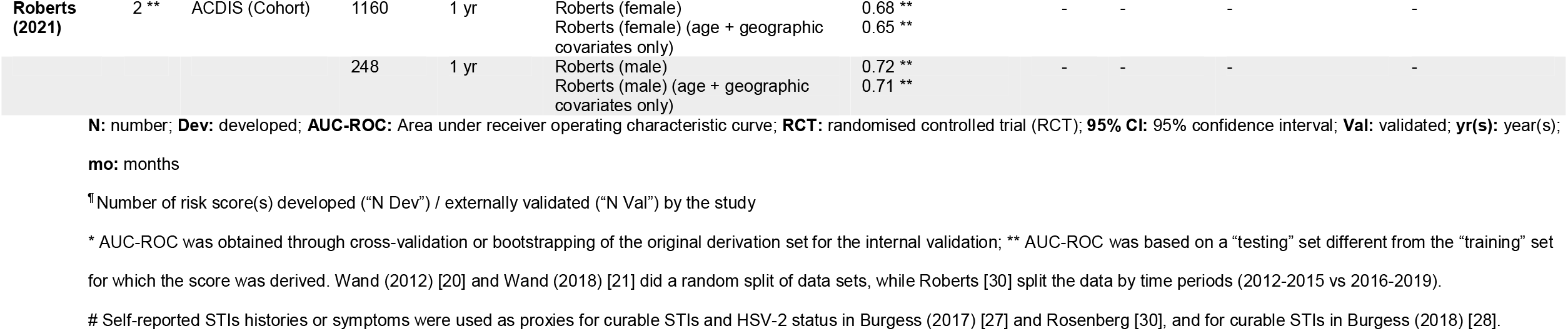
Predictive performance from the development and validation of included risk scores.

When applied to the original data set from which it was developed, the scores had low-to-moderate AUC-ROC ranging from 0.56 to 0.79. Only the VOICE score has been externally validated in other settings. In seven validation studies with AUC-ROC estimates, the accuracy was lower (pooled AUC-ROC: 0.626 [0.588, 0.663]; *I*^*2*^: 64.02%) than in the internal validation (AUC-ROC: 0.69 [0.66, 0.72]) (Table 3, Figure 4). In addition to being among different study populations, it was common for one or two predictors to be missing in external validation sets (Table 3), which may have also contributed to decreased accuracy. Regarding validation of other scores, Roberts developed their risk scores in a large-scale cohort and validated them using data collected in a subsequent time period, also showing moderate discriminatory power (AUC-ROC 0.68 among female and 0.72 among male; Table 3).

**Figure 4.**
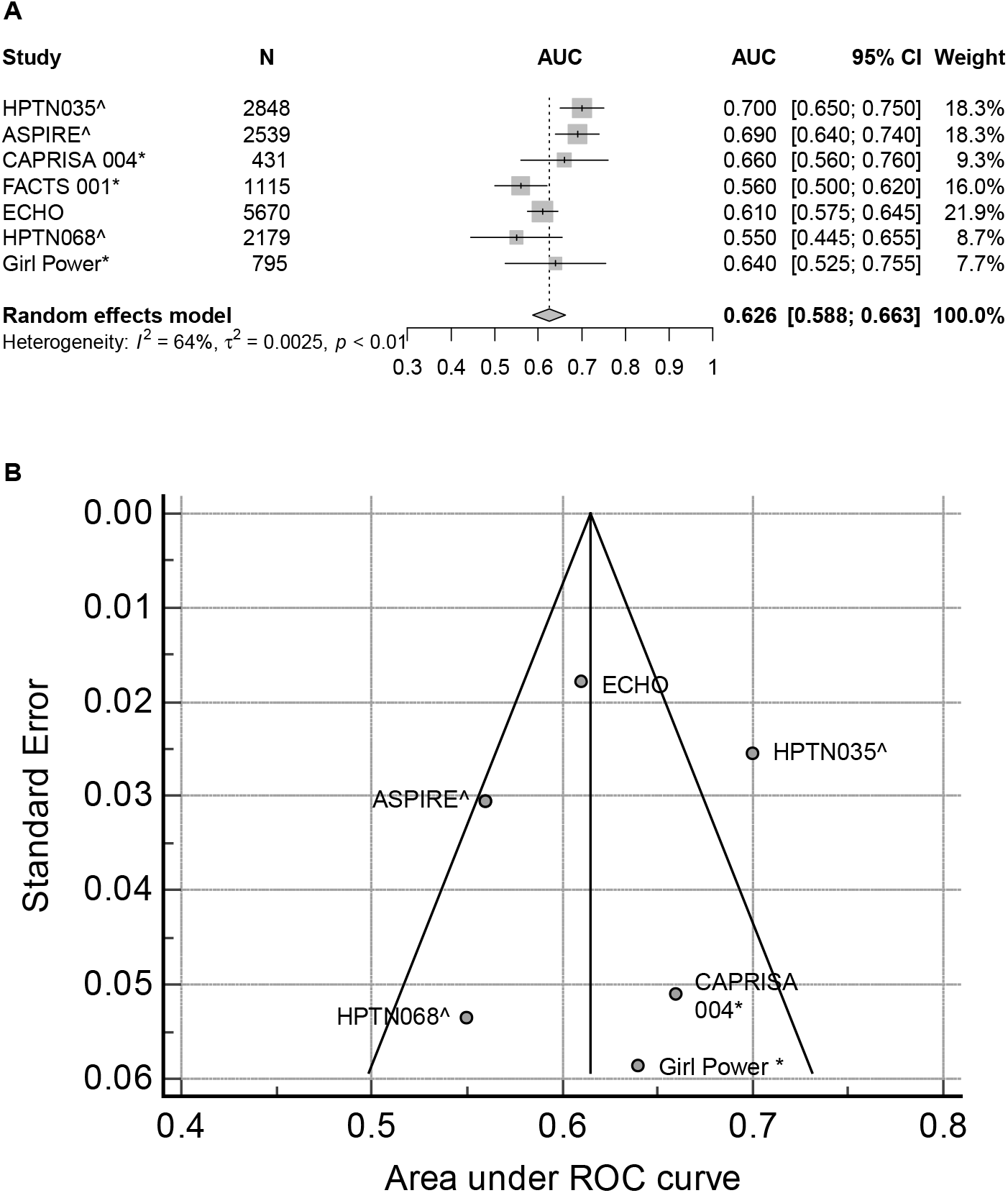
Forest plot (A) and funnel plot (B) for the area-under-curve of the receiver operating characteristic curves (AUC-ROCs) from external validation studies for the VOICE score. Studies with ^^^ did not collect all the predictors intended by the model (Details are in Table 3). Those with ^*^ used self-reported STIs history, syndromic management or self-reported symptoms in place of laboratory diagnosed STIs status at baseline as intended by the original VOICE score (Details are in Table 3 and Table S2). Abbreviations: **AUC:** area under curve; **95% CI:** 95% confidence interval; **ROC:** receiver-operating characteristic.

Several studies compared the discriminative power of combining multivariate risk scores versus single risk factors. Balkus [5] reported that not being married/cohabiting with primary partner alone yielded an AUC-ROC of 0.62 versus 0.69 for the full score, followed by age (0.60) and curable STIs (0.57). In a similar analysis, Peebles^6^ found the most important predictors were age (less than 27), not being married/cohabiting and the provinces of residence. Three studies [8, 9, 28] additionally provided a “modified score” that excluded the laboratory-diagnosed STIs, which are not routinely available in most settings.

Removing laboratory-diagnosed STIs reduced the AUC-ROC by between one to eight percentage-points (Table 3). Roberts found that including only age, HIV prevalence, and viraemia as predictors produced an AUC-ROC of 0.65 for women compared to 0.68 when all risk factors were considered, and 0.71 for men compared to 0.72 when all risk factors were considered (Table 3).

## Incidence among risk group categories

Most studies found that HIV incidence increased monotonically with the risk scores, except for Giovenco [29] (Table S7). Figure 5 shows the proportion of participants identified as high risk compared to the percentage of incident cases contributed by the high-risk group. In six of nine external validation sets of the VOICE score with such information available (Table S7), women with a VOICE score of 5 or above (having around 3 to 4 of the 7 risk factors) had incidence above 3%, the WHO-recommended threshold for PrEP prioritisation. Among studies collecting for all predictors that were intended by the VOICE score (maximum score: 11), above 60% of women scored 5 or above [8, 27, 28]. The threshold for which the observed incidence was >3% varied across populations: in South African samples, Peebles[9] found that incidence was >3% if AGYW scored 3 out of 11, while among the older sample aged 25-34 years only those scoring 6 out of 7 had incidence >3% (16.7% of the sample); in KwaZulu-Natal, Wand [25] observed >3% incidence for 88% of women enrolled in five clinical trials, while in an observational cohort only 60% (3^rd^ quintile and above) of women had incidence >3% [34].

**Figure 5.**
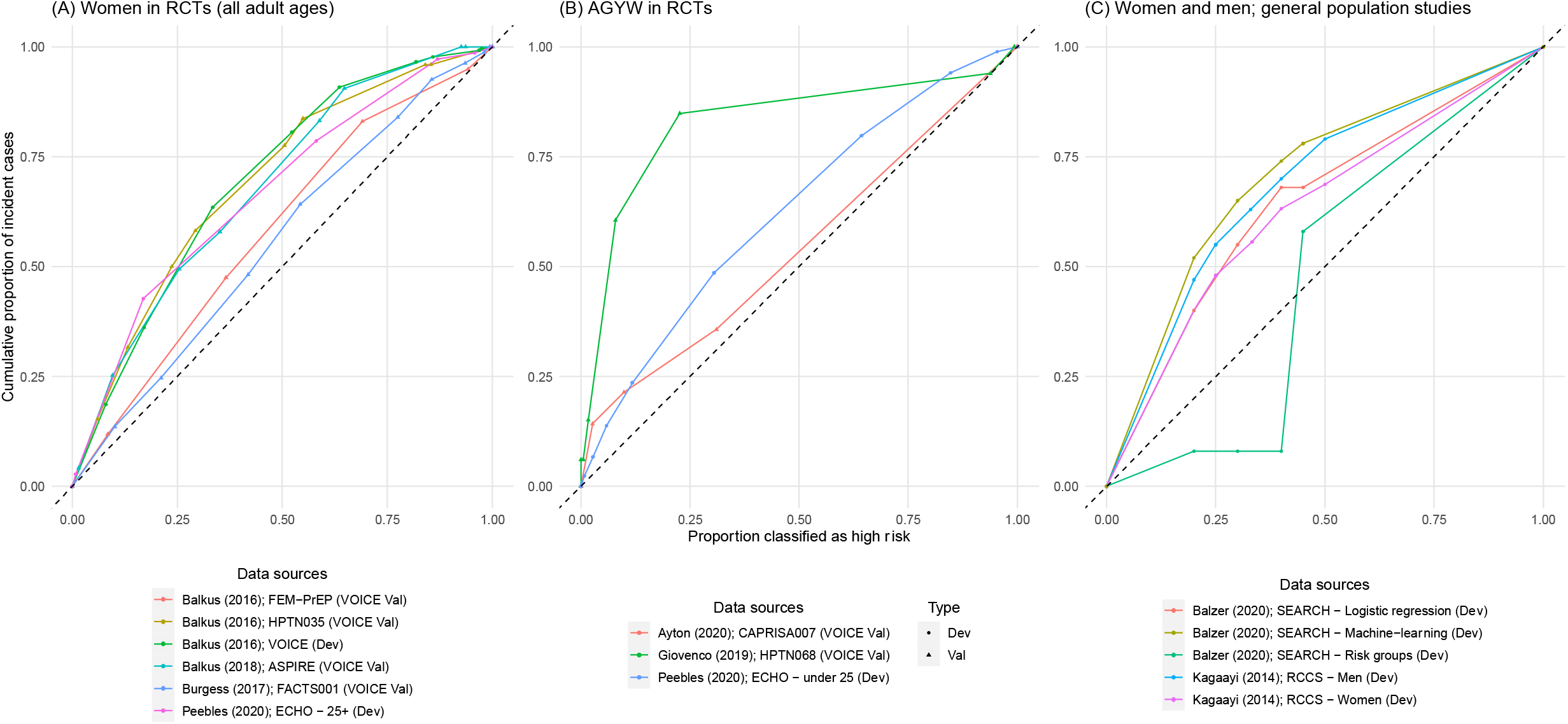
Percentage of individuals identified as high risk among incident cases versus proportion classified as high risk for: (i) women enrolled in clinical trials (A), (ii) AGYW (B), and (iii) the general population (C). When the highest score was used as the threshold, few or none were classified as high risk and they took up a small fraction of all HIV incident cases (indicated by the origin). When the lowest score was used, all were classified as high risk and all incident cases were among them. **Dev**: Development set; **Val:** Validation set. **AGYW:** adolescent girls and young women.

## Discussion

Implementers of HIV programmes in high HIV prevalence settings in sub-Saharan Africa are considering how to optimise HIV prevention, including whether and how to implement HIV risk scoring tools to support identification and prioritisation of persons to receive certain interventions, especially oral PrEP and anticipated future prevention technologies. Our systematic review identified several scoring algorithms developed or validated for this purpose. Risk score development has especially focused on sexually active women of reproductive age or adolescent girls and young women. Twelve of the fifteen sources of studies included data from South Africa and all four risk scores for all-aged women were among RCTs enrolling sexually-active, contraceptive-seeking women in South Africa. Only three studies included men and women [32-34]. Among sexually active women of all ages, younger age, not being married/cohabiting, and having a history of STIs (at baseline or lifetime, both laboratory-confirmed and self-reported) were consistently identified as prognostic factors. Among sexually active AGYW, history of STIs remained consistently selected, but importantly being single/non-cohabiting was not consistently identified. Of the three studies including men, only one reported effect estimates for specific risk factors, with age, education, partner’s occupation, partner’s HIV status, numbers of partners, alcohol before sex, male medical circumcision, STIs, community type, and community HIV prevalence found to be significantly associated with HIV acquisition [32].

Risk scoring based on multiple predictors can improve efficiency in identifying individuals at higher risk of acquiring HIV compared to using individual risk factors [9, 36], but the improvement was only marginal (<0.1 increase in AUC-ROC) [36]. HIV incidence increased steadily with risk score in both development and validation studies, but the ability of risk scores to predict HIV incidence was only moderate. AUC-ROC values ranged from 0.56 to 0.79. AUC-ROC measures the discriminative power of the risk score defined as the probability that a risk score can successfully predict a HIV incident case from a case-and-control pair [37]. An AUC-ROC equal to 1 implies the model perfectly discriminates those who will acquire HIV and those who do not, while 0.5 implies the model has no discriminative power. Most were lower than the AUC-ROC of scores developed for specific populations of sero-discordant couples (AUC-ROC: 0.70 [0.64, 0.76] and 0.76 [0.70,0.83] for two external validation) [38], men-having-sex-with-men in Kenya (0.76 [0.71,0.80]; derivation set) [39], and pregnant and post-partum women in Kenya (0.84 [0.72, 0.95] for derivation set; 0.73 [0.57,0.90] for internal validation set) [40]. The VOICE score was the only model externally validated by multiple studies (nine). Predictive performance of VOICE varied greatly across studies, even among those with all the predictors collected from women seeking contraceptives, which is the original intended population. Among AGYW-only populations, the discriminative power of the VOICE score is expected to be lower because one of the factors, younger age, is fulfilled by everyone in the sample.

Only three of eleven multisite studies considered community-level HIV prevalence or viraemia as a prognostic factor, but all showed it being highly predictive [9, 32, 34]. This supports recommendations to consider both community-level exposure and individual factors to assess individual HIV risk and optimal prevention options. In fact, Roberts found that adding factors beyond community viraemia and age only modestly improved predictive ability, questioning the added value of potentially burdensome screening for more detailed risk behaviours [34]. In contrast, in their analysis adjusted for study sites, Balkus identified non-cohabitation as the most predictive factor, but additional covariates also substantially improved predictions [8]. Further data across multiple settings to adjudicate the added value of more detailed individual risk assessment will help guide HIV programme implementation strategies.

The implication of the only moderate discrimination is that any use of risk scores to determine eligibility for certain prevention modalities will either restrict access for a large share of individuals who are at risk for future infection or require either setting a very low threshold score to ensure a high proportion of infections are included. In the latter case, the burden of implementing the screening tool may not outweigh the benefit, if only a relatively small share of the population are ultimately screened out. Rather than restricting eligibility, another potential use of risk scores may be as a tool to prompt discussion about HIV prevention to individuals or in settings where it might otherwise not be offered. The consistently identified risk factors offer some promise this could be valuable. The threshold for such an offer could be differentiated according to local context: a relatively high threshold in settings with low community prevalence or viraemia and a lower threshold in areas with higher community exposure.

### Discriminatory ability of risk scores

There are several possible reasons risk scores based on well-established risk factors are only moderately discriminative. First, HIV risk can change rapidly over short time intervals with life course events. Risk assessed at baseline may only be moderately predictive of an individual’s actual HIV risk six to twelve months later. Individuals identified as low risk at baseline may become high-risk over the time due to changes in their behaviours, their partners’ behaviours, or migration into new communities. Second, risk of acquiring HIV depends not only on individual-level risk factors but also predictors related to their partners and communities. Consequently, adults with behaviour considered ‘low risk’, such as a single cohabiting sex partner, could still be exposed to high risk of HIV infection if their partner acquires HIV. While the HIV incidence rate among this group is relatively low, they may contribute a large proportion of total new infections, fundamentally limiting the extent to which HIV prevention can be optimised without specific, timely, and accurate information about risk among sexual partners. Both the number of partners and partner having other partners were significantly associated with HIV acquisition in around half of the reported risk scores (Table 2). Third, factors included in risk scores are susceptible to reporting or measurement errors to varying degrees.

Recent STI, identified through laboratory diagnosis in the clinical trials used for risk score development, was the most consistently identified predictive factor for HIV infection.

However, laboratory testing for STIs is not routinely available in most low- and middle-income countries, where they were typically diagnosed through syndromic management instead. In our review, validation studies using self-reported or syndromic identified STIs [27, 28, 30] had similar accuracy (AUC-ROCs) as those using laboratory tests [9] (Table 3), but elsewhere syndromic management has consistently had only low to moderate accuracy [41-43].

### Limited generalisability of risk scores

Generalising and applying the risk scores reviewed here across high burden settings faces several challenges. First, data were disproportionately from South Africa, which has unique HIV epidemiology and low rates of marriage and cohabitation compared to neighbouring countries. Risk factors consistently identified to be significant for sexually active, contraceptive-seeking women (younger age, non-cohabitation and STIs), were all from South African studies. These may not generalise to other settings with higher marriage rates and younger age at marriage.

Second, some data used to develop and validate risk scores were relatively old, with about half of studies completed before 2012 when HIV incidence was higher and ART coverage lower. Rapid scale-up of ART, commensurate changes in community-level unsuppressed viral load, and shifting distribution of new infections to older ages have affected exposure to HIV infection, and consequently risk associated with individual characteristics may have changed over time and vary across settings. Considering how transmission dynamics interact with identified risk factors will be important to ensure context appropriate focusing of HIV prevention in a continually evolving epidemic [44].

Third, seven out of thirteen studies focused on the sexually active, contraceptive-seeking women enrolled in RCTs, who were intentionally selected as relatively high-risk for testing novel HIV prevention technologies. They excluded those who did not attend STI or family clinics (for studies based on clinical sites) and who intended to be pregnant within one or two years. External validation of the VOICE score by Giovenco (2019) demonstrated that the score did not generalise to school-attending AGYW, a majority of whom were young and not cohabiting with a primary partner, but also not sexually active at baseline assessment [29].

Fourth, there were subtle differences in definition and coding of risk factors across studies. This undermines the appropriateness of our pooled risk ratio estimates. In many validation studies, some selected risk factors were not available or defined differently [26, 29, 30].

Inconsistencies in defining and measuring certain risk factors like the partnership and behavioural factors may have resulted in some important but inconsistently reported predictors being overlooked.

### Methodological challenges

More generally, developing risk scores for HIV incidence is fundamentally challenging, resulting in moderate to high assessed risk of bias using the PROBAST checklist (Table S3). As HIV infection is a relatively uncommon event, in most studies the ratio of cases observed to risk factors considered was far lower than recommended. Many studies were limited in accounting for over-fitting and model optimism, clarity about handling missing data [16]. Our review was also constrained by incomplete reporting of multivariate regression results of initial and final models in some studies. Only a few studies compared the AUC-ROC of the full models with that of individual predictors, making it difficult to draw conclusions about the necessity of detailed risk assessment compared to a few key characteristics—a key question for HIV programme implementation. Finally, we only focused on the heterosexual adult population and did not consider risk scores among key and vulnerable populations with high incidence. Other epidemiological evidence strongly supports prioritisation and provision of HIV prevention for these groups where they can be identified.

### Future research priorities

Our review identified three priorities for future studies. First, comparison of the AUC-ROCs of the full model versus individual predictors or more parsimonious models will help differentiate key predictors for identifying risk groups and prioritising resources and the relative value of factors that are more invasive or intensive to collect. Use of machine-learning techniques has also showed a potential to improving prediction accuracy and can be incorporated into some prevention interventions [45].

Second, additional risk score development and validation using recent incidence data from wider geographic settings will increase the generalisability of HIV risk scores. Finally, although in our review all AUC-ROCs in the external validation studies fell below 0.7, classified as poor discrimination by some [37], the discrimination of the risk scores may be higher when applied outside selected RCT populations that include not sexually individuals who would likely be screened out by risk scores, but were systematically excluded from the study populations. Alternately, individuals not sexually active at a baseline risk assessment, but who become active, could be an important risk population missed by the studies in our review. This could be explored through modelling, and further extended to study the infections averted, resources saved, and cost effectiveness of incorporating multivariable risk scores into risk stratification and prevention strategy prioritisation, and to model counterfactuals incidence for active control trials and implementation studies [44]. Our findings inform such analyses by providing data on the incidence rate ratios and proportion of infections among each group compared to the size of the group.

## Conclusions

Several risk scores have been developed for identifying individuals at increased risk for HIV among general populations in sub-Saharan Africa. Among sexually active, contraceptive-seeking women, these studies have consistently identified younger age, not being married/cohabiting and STIs as risk factors. These consistently identified risk factors may be useful to prompt discussions or offers of efficacious HIV prevention. However, taken together, the programmatic benefit of implementing HIV risk scores as screening or triaging tools may be limited due to only moderate overall discriminatory ability and limited improvement compared to focusing on geographic areas with high HIV burden and basic demographics such as age group. The marginal benefits must be balanced with additional administrative burden for providers and consideration for whether screening questions could be perceived as stigmatizing, invasive, or exclusionary for clients.

## Data Availability

The manuscript is a systematic review. All extracted data are reported in the supplementary material.

## Authors’ contributions

KMJ and JWE conceptualised th review. KMJ did the initial literature search and wrote the protocol with substantial inputs from JWE. AH, HE, OE, KMJ, KL, and MT screened abstracts and full tests, extracted the data, and performed critical appraisal. KMJ performed the analysis and wrote the first draft of the manuscript with substantial inputs from JWE, OE and AH. KMJ, JWE, AH, HE, OE, KL, and MT contributed to interpretation of the results and edited the manuscript for intellectual content. All authors read and approved the final version of the manuscript.

## Acknowledgments

This research was supported by the Bill & Melinda Gates Foundation (Grant numbers: OPP1190661, INV-002606, OPP1164897), UNAIDS, and the MRC Centre for Global Infectious Disease Analysis (reference MR/R015600/1), jointly funded by the UK Medical Research Council (MRC) and the UK Foreign, Commonwealth & Development Office (FCDO), under the MRC/FCDO Concordat agreement and is also part of the EDCTP2 programme supported by the European Union.

We thank Natsuko Imai for providing technical guidance and support, Adam Akullian and Allen Roberts for providing useful comments and unpublished details of their study in this review.

## Additional files

Additional file 1: Risk scores for predicting HIV incidence among general population in sub-Saharan Africa: a systematic review and meta-analysis—Supplementary Materials

File format: PDF

Description: This supplementary materials contain: (i) details of search and data extraction strategies, (ii) detailed characteristics of the data sources, (iii) summary of risk of bias assessment, (iv) summary of additional analyses of study results, and (v) PRISMA checklists.

## List of abbreviations

AGYW: adolescent girls and young women
(a)HR: (adjusted) hazard-ratio
AUC-ROC: area under the receiver operating characteristic curve
CHARMS: Critical Appraisal and Data Extraction for Systematic Reviews of Prediction Modelling Studies Checklist
HSV-2: Human Simplex Virus – 2
MMC: male medical circumcision
PrEP: pre-exposure prophylaxis
PROBAST: Prediction Model Risk of Bias assessment tool checklist
PRISMA: Preferred Reporting Items for Systematic Reviews and Meta-Analyses
RCT: randomised controlled trial
STI: sexually transmitted infection

