## Supplementary Information for "Risk scores for predicting HIV incidence among adult heterosexual populations in sub-Saharan Africa: a systematic review and meta-analysis"

#### Appendix I Database search strategy
**Search date: 15th February 2021 (Monday)**

##### 1. MEDLINE via Ovid

| | Query |
|---|---|
| **1** | ((risk* adj appraisal*) or (risk* adj algorithm*) or (risk* adj “assessment tool”) or (risk* adj1 calculat*) or (risk* adj chart*) or (risk* adj1 checklist*) or (risk* adj “classification tool”) or (risk* adj disk) or (risk* adj disc?) or (risk* adj function*) or (risk* adj equation*) or (risk* adj1 index) or (risk* adj1 indices) or (risk* adj3 scale*) or (risk* adj3 scor*) or (risk* adj “stratification tool”) or (risk* adj table*) or (risk* adj threshold*) or (risk* adj3 tool*) or (risk* adj prediction*) or ("risk assessment" adj function*) or (prognostic adj tool) or (prognostic adj model) or ((risk or inciden* or hazard* or prognos*) and ((scor* adj algorithm*) or (scor* adj scheme*) or (scor* adj system*) or (scor* adj tool*) or (screening adj score*) or (prediction adj equation) or (predicti* adj instrument*) or (predicti* adj model*) or (predicti* adj rule) or (predicti* adj scor*) or (projecti* adj1 risk*)))).tw |
| **2** | exp decision support techniques/ or exp clinical decision rules/ or exp data interpretation, statistical/ [included all subheadings] |
| **3** | exp Nomograms/ |
| **4** | OR # 2-#3 |
| **5** | exp algorithms/ or exp artificial intelligence/ or exp latent class analysis/ |
| **6** | (screen* or scor* or predict* or risk* or prognos*).tw. |
| **7** | AND #5 - #6 |
| **8** | OR #1, #4, #7 |
| **9** | (Africa* or Angola or Benin or Botswana or “Burkina Faso” or Burundi or “Cabo Verde” or “Cape Verde” or Cameroon or “Central African Republic” or Chad or Comoros or “Democratic Republic of Congo” or DRC or “Republic of Congo” or “Cote d'Ivoire” or “Cote D' Ivoire” or “Ivory Coast” or Djibouti or “Equatorial Guinea” or Eritrea or Eswatini or Ethiopia or Gabon or Gambia or Ghana or Guinea or Guinea-Bissau or Kenya or Lesotho or Liberia or Madagascar or Malawi or Mali or Mauritania or Mauritius or Mozambique or Namibia or Niger or Nigeria or Rwanda or (“Sao Tome” adj1 Princip*) or Senegal or Seychelles or “Sierra Leone” or Somalia or “South Africa” or “South Sudan” or Sudan or Swaziland or Tanzania or Togo or Uganda or Zambia or Zimbabwe).mp |
| **10** | (HIV* or “human immunodeficiency virus” or “human-immunodeficiency-virus” or AID* or “Acquired Immunodeficiency Syndrome”).ti. |
| **11** | AND # 8 - #10 |

##### 2. Embase, MIDIRS, APA PsycInfo, Global Health via Ovid

| | Query |
|---|---|
| **1** | ((risk* adj appraisal*) or (risk* adj algorithm*) or (risk* adj “assessment tool”) or (risk* adj1 calculat*) or (risk* adj chart*) or (risk* adj1 checklist*) or (risk* adj “classification tool”) or (risk* adj disk) or (risk* adj disc?) or (risk* adj function*) or (risk* adj equation*) or (risk* adj1 index) or (risk* adj1 indices) or (risk* adj3 scale*) or (risk* adj3 scor*) or (risk* adj “stratification tool”) or (risk* adj table*) or (risk* adj threshold*) or (risk* adj3 tool*) or (risk* adj prediction*) or ("risk assessment" adj function*) or (prognostic adj tool) or (prognostic adj model) or ((risk or inciden* or hazard* or prognos*) and ((scor* adj algorithm*) or (scor* adj scheme*) or (scor* adj system*) or (scor* adj tool*) or (screening adj score*) or (prediction adj equation) or (predicti* adj instrument*) or (predicti* adj model*) or (predicti* adj rule) or (predicti* adj scor*) or (projecti* adj1 risk*)))).tw |
| **2** | (Africa* or Angola or Benin or Botswana or “Burkina Faso” or Burundi or “Cabo Verde” or “Cape Verde” or Cameroon or “Central African Republic” or Chad or Comoros or “Democratic Republic of Congo” or DRC or “Republic of Congo” or “Cote d'Ivoire” or “Cote D' Ivoire” or “Ivory Coast” or Djibouti or “Equatorial Guinea” or Eritrea or Eswatini or Ethiopia or Gabon or Gambia or Ghana or Guinea or Guinea-Bissau or Kenya or Lesotho or Liberia or Madagascar or Malawi or Mali or Mauritania or Mauritius or Mozambique or Namibia or Niger or Nigeria or Rwanda or (“Sao Tome” adj1 Princip*) or Senegal or Seychelles or “Sierra Leone” or Somalia or “South Africa” or “South Sudan” or Sudan or Swaziland or Tanzania or Togo or Uganda or Zambia or Zimbabwe).mp |
| **3** | (HIV* or “human immunodeficiency virus” or “human-immunodeficiency-virus” or AID* or “Acquired Immunodeficiency Syndrome”).ti. |
| **4** | AND # 1 - #3 |
| **5** | remove duplicates from # 4 |

**3. Scopus**

| | Query |
|---|---|
| **1** | TITLE-ABS-KEY ((risk* W/0 appraisal*) or (risk* W/0 algorithm*) or (risk* W/0 “assessment tool”) or (risk* W/1 calculat*) or (risk* W/0 chart*) or (risk* W/1 checklist*) or (risk* W/0 “classification tool”) or (risk* W/0 disk) or (risk* W/0 disc?) or (risk* W/0 function*) or (risk* W/0 equation*) or (risk* W/1 index) or (risk* W/1 indices) or (risk* W/3 scale*) or (risk* W/3 scor*) or (risk* W/0 “stratification tool”) or (risk* W/0 table*) or (risk* W/0 threshold*) or (risk* W/3 tool*) or (risk* W/0 prediction*) or ("risk assessment" W/0 function*) or (prognostic W/0 tool) or (prognostic W/0 model) or ((risk OR inciden* OR hazard* OR prognos*) and ((scor* W/0 algorithm*) or (scor* W/0 scheme*) or (scor* W/0 system*) or (scor* W/0 tool*) or (screening W/0 score*) or (prediction W/0 equation) or (predicti* W/0 instrument*) or (predicti* W/0 model*) or (predicti* W/0 rule) or (predicti* W/0 scor*) or (projecti* W/0 risk*)))) |
| **2** | ALL (Africa* or Angola or Benin or Botswana or “Burkina Faso” or Burundi or “Cabo Verde” or “Cape Verde” or Cameroon or “Central African Republic” or Chad or Comoros or “Democratic Republic of Congo” or DRC or “Republic of Congo” or “Cote d'Ivoire” or “Cote D' Ivoire” or “Ivory Coast” or Djibouti or “Equatorial Guinea” or Eritrea or Eswatini or Ethiopia or Gabon or Gambia or Ghana or Guinea or Guinea-Bissau or Kenya or Lesotho or Liberia or Madagascar or Malawi or Mali or Mauritania or Mauritius or Mozambique or Namibia or Niger or Nigeria or Rwanda or (“Sao Tome” W/1 Princip*) or Senegal or Seychelles or “Sierra Leone” or Somalia or “South Africa” or “South Sudan” or Sudan or Swaziland or Tanzania or Togo or Uganda or Zambia or Zimbabwe) |
| **3** | TITLE(HIV* or "human immunodeficiency virus" or "human-immunodeficiency-virus" or AID* or "Acquired Immunodeficiency Syndrome") |
| **4** | AND # 1 - #3 |

**4. CINAHL (EBSCO)**

| | Query |
|---|---|
| **1** | Title, Abstract, Subject Headings and Keywords:<br>((risk* N0 appraisal*) or (risk* N0 algorithm*) or (risk* N0 “assessment tool”) or (risk* N1 calculat*) or (risk* N0 chart*) or (risk* N1 checklist*) or (risk* N0 “classification tool”) or (risk* N0 disk) or (risk* N0 disc?) or (risk* N0 function*) or (risk* N0 equation*) or (risk* N1 index) or (risk* N1 indices) or (risk* N3 scale*) or (risk* N3 scor*) or (risk* N0 “stratification tool”) or (risk* N0 table*) or (risk* N0 threshold*) or (risk* N3 tool*) or (risk* N0 prediction*) or ("risk assessment" N0 function*) or (prognostic N0 tool) or (prognostic N0 model) or ((risk OR inciden* OR hazard* OR prognos*) AND ((scor* N0 algorithm*) or (scor* N0 scheme*) or (scor* N0 system*) or (scor* N0 tool*) or (screening N0 score*) or (prediction N0 equation) or (predicti* N0 instrument*) or (predicti* N0 model*) or (predicti* N0 rule) or (predicti* N0 scor*) or (projecti* N0 risk*)))) |
| **2** | (MH "Clinical Prediction Rules") |
| **3** | 1 OR 2 |
| **4** | All texts:<br>(Africa* or Angola or Benin or Botswana or “Burkina Faso” or Burundi or “Cabo Verde” or “Cape Verde” or Cameroon or “Central African Republic” or Chad or Comoros or “Democratic Republic of Congo” or DRC or “Republic of Congo” or “Cote d'Ivoire” or “Cote D' Ivoire” or “Ivory Coast” or Djibouti or “Equatorial Guinea” or Eritrea or Eswatini or Ethiopia or Gabon or Gambia or Ghana or Guinea or Guinea-Bissau or Kenya or Lesotho or Liberia or Madagascar or Malawi or Mali or Mauritania or Mauritius or Mozambique or Namibia or Niger or Nigeria or Rwanda or (“Sao Tome” N1 Princip*) or Senegal or Seychelles or “Sierra Leone” or Somalia or “South Africa” or “South Sudan” or Sudan or Swaziland or Tanzania or Togo or Uganda or Zambia or Zimbabwe) |
| **6** | Title<br>HIV* or "human immunodeficiency virus" or "human-immunodeficiency-virus" or AID* or "Acquired Immunodeficiency Syndrome" |
| **7** | AND # 3 - # 6 |

#### 5. Cochrane

| | Query |
|---|---|
| **1** | Title / Abstract / Keyword:<br>((risk* NEXT appraisal*) or (risk* NEXT algorithm*) or (risk* NEXT “assessment tool”) or (risk* NEAR/1 calculat*) or (risk* NEXT chart*) or (risk* NEAR/1 checklist*) or (risk* NEXT “classification tool”) or (risk* NEXT disk) or (risk* NEXT disc?) or (risk* NEXT function*) or (risk* NEXT equation*) or (risk* NEAR/1 index) or (risk* NEAR/1 indices) or (risk* NEAR/3 scale*) or (risk* NEAR/3 scor*) or (risk* NEXT “stratification tool”) or (risk* NEXT table*) or (risk* NEXT threshold*) or (risk* NEAR/3 tool*) or (risk* NEXT prediction*) or ("risk assessment" NEXT function*) or (prognostic NEXT tool) or (prognostic NEXT model) or ((risk OR inciden* OR hazard* OR prognos*) and ((scor* NEXT algorithm*) or (scor* NEXT scheme*) or (scor* NEXT system*) or (scor* NEXT tool*) or (screening NEXT score*) or (prediction NEXT equation) or (predicti* NEXT instrument*) or (predicti* NEXT model*) or (predicti* NEXT rule) or (predicti* NEXT scor*) or (projecti* NEXT risk*)))) |
| **2** | All text:<br>(Africa* or Angola or Benin or Botswana or “Burkina Faso” or Burundi or “Cabo Verde” or “Cape Verde” or Cameroon or “Central African Republic” or Chad or Comoros or “Democratic Republic of Congo” or DRC or “Republic of Congo” or “Cote d'Ivoire” or “Cote D' Ivoire” or “Ivory Coast” or Djibouti or “Equatorial Guinea” or Eritrea or Eswatini or Ethiopia or Gabon or Gambia or Ghana or Guinea or Guinea-Bissau or Kenya or Lesotho or Liberia or Madagascar or Malawi or Mali or Mauritania or Mauritius or Mozambique or Namibia or Niger or Nigeria or Rwanda or (“Sao Tome” NEAR/1 Princip*) or Senegal or Seychelles or “Sierra Leone” or Somalia or “South Africa” or “South Sudan” or Sudan or Swaziland or Tanzania or Togo or Uganda or Zambia or Zimbabwe) |
| **3** | Record title:<br>(HIV* or "human immunodeficiency virus" or "human-immunodeficiency-virus" or AID* or "Acquired Immunodeficiency Syndrome") |
| **4** | 1 AND 2 AND 3 |

#### 6. Web of Science

| | Query |
|---|---|
| **1** | TS = ((risk* NEAR/0 appraisal*) or (risk* NEAR/0 algorithm*) or (risk* NEAR/0 “assessment tool”) or (risk* NEAR/1 calculat*) or (risk* NEAR/0 chart*) or (risk* NEAR/1 checklist*) or (risk* NEAR/0 “classification tool”) or (risk* NEAR/0 disk) or (risk* NEAR/0 disc?) or (risk* NEAR/0 function*) or (risk* NEAR/0 equation*) or (risk* NEAR/1 index) or (risk* NEAR/1 indices) or (risk* NEAR/3 scale*) or (risk* NEAR/3 scor*) or (risk* NEAR/0 “stratification tool”) or (risk* NEAR/0 table*) or (risk* NEAR/0 threshold*) or (risk* NEAR/3 tool*) or (risk* NEAR/0 prediction*) or ("risk assessment" NEAR/0 function*) or (prognostic NEAR/0 tool) or (prognostic NEAR/0 model) or ((risk OR inciden* OR hazard* OR prognos*) and ((scor* NEAR/0 algorithm*) or (scor* NEAR/0 scheme*) or (scor* NEAR/0 system*) or (scor* NEAR/0 tool*) or (screening NEAR/0 score*) or (prediction NEAR/0 equation) or (predicti* NEAR/0 instrument*) or (predicti* NEAR/0 model*) or (predicti* NEAR/0 rule) or (predicti* NEAR/0 scor*) or (projecti* NEAR/0 risk*)))) |
| **2** | ALL = (Africa* or Angola or Benin or Botswana or “Burkina Faso” or Burundi or “Cabo Verde” or “Cape Verde” or Cameroon or “Central African Republic” or Chad or Comoros or “Democratic Republic of Congo” or DRC or “Republic of Congo” or “Cote d'Ivoire” or “Cote D' Ivoire” or “Ivory Coast” or Djibouti or “Equatorial Guinea” or Eritrea or Eswatini or Ethiopia or Gabon or Gambia or Ghana or Guinea or Guinea-Bissau or Kenya or Lesotho or Liberia) |
| **3** | ALL = (Madagascar or Malawi or Mali or Mauritania or Mauritius or Mozambique or Namibia or Niger or Nigeria or Rwanda or (“Sao Tome” AND Princip*) or Senegal or Seychelles or “Sierra Leone” or Somalia or “South Africa” or “South Sudan” or Sudan or Swaziland or Tanzania or Togo or Uganda or Zambia or Zimbabwe) |
| **4** | OR #2 - #3 |
| **5** | TI = (HIV* or "human immunodeficiency virus" or "human-immunodeficiency-virus" or AID* or "Acquired Immunodeficiency Syndrome") |
| **6** | AND # 1, #4, #5 |

#### Appendix II. Details on data extraction

The following items are extracted from the included studies:

| | |
|---|---|
| **Study** | 1. Study design<br>2. Study period<br>3. Countries of study |
| **Participants** | 1. Number and location of sites,<br>2. Inclusion/exclusion criteria<br>3. Participant description<br>4. Intervention(s) received (if any) and effectiveness |
| **Sample size** | 1. Number of participants enrolled<br>2. Number of participants analysed<br>3. Reason(s) for exclusion of enrolled participants from analysis<br>4. Number of HIV incident cases<br>5. Number of HIV incident cases per candidate predictor (Events Per Variable) |
| **Missing data** | 1. Number of participants with any missing values (predictors)<br>2. Loss-to-follow-up (i.e., absence of at least one follow-up HIV test)<br>3. Method for handling missing data |
| **Outcomes** | 1. Definition and method for measurement of outcome<br>2. Was the same outcome definition (and method for measurement) used in all patients?<br>3. Type of outcome (e.g., single or combined endpoints)<br>4. Was the outcome assessed without knowledge of the candidate predictors (i.e., blinded)?<br>5. Were candidate predictors part of the outcome (e.g., in panel or consensus diagnosis)?<br>6. Time point where the outcome is determined<br>7. Total duration of follow-up<br>8. HIV incidence |
| **Predictors** | 1. Timing of predictor measurement<br>2. Number and type of predictors<br>3. Methods for measurement of sexually transmitted infections (STIs)<br>4. Were predictors assessed blinded for outcome, and for each other (if relevant)?<br>5. Handling of predictors in the modelling (e.g., continuous, linear,<br>6. non-linear transformations or categorised) |
| **Model development** | 1. Modelling approach<br>2. Modelling assumptions satisfied<br>3. Method for selection of predictors for inclusion in multivariable modelling<br>4. Method for selection of predictors during multivariable modelling<br>5. Shrinkage of predictor weights or regression coefficients |
| **Model performance assessment** | 1. Calibration and discrimination measures that have been used<br>2. Classification measures and how cut points have been determined |
| **Model evaluation** | 1. Method used for testing model performance: internal validation (e.g., bootstrapping, cross-validation or none) or external validation<br>2. In case of poor validation, whether model was adjusted or updated |
| **Results** | 1. Predictors retained in the final and other multivariable models (e.g., excluding the laboratory diagnosed STIs)<br>2. Coefficients for the predictors (i.e., effect size estimates)<br>3. Model performance measures (with confidence intervals)<br>4. Comparison of the distribution of predictors (including missing data) for development and validation datasets |

#### Table S1. Characteristics of the included studies

| First author (Year); | Any intervention; if yes, is the intervention effective and adjusted in the model? | Description of dataset; Inclusion / exclusion criteria | Age | Timing of outcome determination | Candidate predictors | Predictors selected for inclusion into the model | Predictors retained in the final model |
|---|---|---|---|---|---|---|---|
| **(I) Women only (All ages / 25+years old)** | | | | | | | |
| **Wand (2012) [1]** | **MIRA** [2]<br>**Intervention**:<br>Latex diaphragm, lubricant gel, and condoms (intervention) vs condoms alone (control)<br><br>**Outcome:**<br>Prevention of heterosexual HIV acquisition among women<br><br>**Any significant effect(s):**<br>No. The risk score was developed by using the Durban data only. The intervention did not show a significant effect in reducing HIV incidence in the Durban sites (Hazard Ratio [HR]: 0.95 [0.69, 1.31]).<br><br>**Intervention incorporated as a predictor in the model?**<br>No. Only contraception uses at baseline and condom use in the past 3 months before enrolment were included. | Randomised controlled trial (RCT);<br><br>(i) sexually active women,<br><br>(ii) willing to use contraception / not planning to get pregnant in the next 24 months from two sites,<br><br>(iii) residing in Durban, South Africa | **Mean**: 27;<br><br>**IQR**: 22-34 | **Outcome determination**: Quarterly followed-up with a total period ranging from 12 to 24 months<br><br>**Prediction horizon:**<br>any event during the entire follow-up period | • Age<br>• Cohabitation status (whether the participant was living with her sexual partner)<br>• Level of education<br>• Employment status<br>• Number of lifetime sexual partners<br>• Age at first sex<br>• Consistent condom use (in past three months)<br>• Contraception use [long term (tubal ligation, vasectomy, intrauterine device), hormonal injectables, oral contraceptives), barrier methods (male/female condoms)]<br>• Average number of weekly sex acts<br>• Partner risk (defined as one or more of: any sexual partners testing positive for HIV;<br>• suspecting or knowing that their regular partner had other sex partners in the past three months; any vaginal sex when partner was under influence of drugs or alcohol in past three months; regular partner was away from home for one or more months)<br>• High behavioural risk (defined as one or more of: any exchange of sex for money, food, drugs, or shelter; two or more sexual partners within past three months; ever having vaginal sex under influence of drugs or alcohol in past three months; ever using a needle for injectable drug use; having anal sex in past three months)<br>• Any STIs (Neisseria gonorrhoeae, Chlamydia<br>• trachomatis and syphilis)<br>• Herpes simplex virus<br>• 2 (HSV-2) in the past six months<br>• Any reproductive tract infections (candidiasis, bacterial vaginosis),<br>• Genital epithelial disruption<br>• Genital signs<br>• Genital discharge<br>• Genital ulcer<br>• Abnormal vulva | **Criteria for variable to be included**: univariate association (P threshold not mentioned)<br><br>• Age (<25, 25-34, 35+ yrs)<br>• Lifetime male sexual partners<br>• Behaviour risk<br>• Coital frequency (per week)<br><br>• Cohabiting with a sex partner<br>• Genital epithelial disruption<br>• Genital signs<br>• Genital discharge<br>• Genital Ulcer<br>• Abnormal vulva | **Cox regression**;<br>Stepwise backward elimination procedures<br><br>**Final model:**<br>• Lifetime male sexual partners<br>• Behavioural risk<br>• Cohabiting with a sexual partner<br>• Genital epithelial disruption<br>• Genital discharge |
| **Wand (2018) [3]** | The risk score was developed based on KwaZulu Natal, South Africa only. At all visits, the participants received counselling on risk reduction and had access to male condoms as desired [3].<br><br>**MIRA**[2]<br>**Intervention**:<br>Latex diaphragm, lubricant gel, and condoms (intervention) vs condoms alone (control)<br><br>**Outcome:**<br>Prevention of heterosexual HIV acquisition among women<br><br>**Any significant effect(s):**<br>No. The intervention did not show a significant effect in reducing HIV incidence in the KZN (Durban) sites (adjusted HR: 0.95 [0.69, 1.31]). | RCTs;<br><br>(i) sexually active women,<br><br>(ii) willing to use contraception / not planning to get pregnant,<br><br>(iii) residing in KwaZulu Natal, South Africa. | **Median** 27; **IQR:** 22–33 | **Outcome determination**: Various timepoint depending on studies<br><br>**Prediction horizon:**<br>any event during the entire follow-up period | • Age (<20, 20–24, 25–29, 30–34, 35–39 and 40+ years);<br>• Married/cohabiting with a sexual partner (yes/no)<br>• Level of education (less than high school vs. completed high school or above)<br>• Number of sexual partners in past three months (3+ vs.<3);<br>• age at sexual debut (<16 vs. 16+ years),<br>• condom used at last sex (yes/no).<br>• Injectables (yes/no),<br>• oral contraceptives (pills) (yes/no)<br>• male condom (yes/no);<br>• parity (null/primiparity: <2 births, multi-parity: 2 vs. multi-parity: 3+)<br>• diagnosed with STIs (chlamydia, gonorrhoea, syphilis)<br>• language spoken at home,<br>• employment/regular income (yes vs. no),<br>• partner's circumcision status<br>• average number of sexual acts in the past 7 days.) | **Predictors with univariate association P<0.05 were included.**<br><br>**Variables included:**<br>• Age<br>• Age at sexual debut<br>• education,<br>• employment,<br>• partner's circumcision status,<br>• condom not used in last sex<br>• Married/cohabiting with a sexual partner<br>• Number of sexual partners<br>• Parity<br>• Injectable contraception<br>• Diagnosed with STIs | **Cox regression**.<br>Stepwise backward elimination procedures<br><br>**Final model:**<br>• Age<br>• Age at sexual debut<br>• Married/cohabiting with a sexual partner<br>• Number of sexual partners<br>• Parity<br>• Injectable contraception<br>• Diagnosed with STIs |

| | | | |
|---|---|---|---|
| | **MDP 301 [4]**<br>**Intervention:**<br>2% PRO2000, 0.5% PRO2000, or placebo gel groups.<br><br>At the 12, 24, 40 and 52 week clinic visits, women were provided with HIV testing and counselling with promotion of safer sex practices, provision of free condoms, and diagnosis and treatment of sexually transmitted diseases.<br><br>**Outcome:**<br>Prevention of HIV incidence<br><br>**Any significant effect(s):**<br>No. HIV incidence was much the same between groups overall (HR for 0.5% PRO2000 vs placebo: 1.05 [0.82; 1.34], p=0.71). | | • Education<br>• Employment<br>• Partner's circumcision status<br>• Condom not used in last sex |
| | **NCT00213083 [5]**<br>**Intervention:**<br>Microbicide gel (Carraguard) plus condoms vs placebo gel plus condoms.<br><br>**Outcome:**<br>Prevention of HIV incidence<br><br>**Any significant effect(s):**<br>No. HIV was not significantly different between groups (adjusted HR: 0.87 [0.69; 1.09]) overall. | | |
| | **VOICE [6]**<br>**Interventions:**<br>5 arms: (i) oral TDF (300 mg) and TDF-FTC placebo, (ii) oral TDF-FTC (300 mg of TDF and 200 mg of FTC) and TDF placebo, (iii) oral TDF placebo and oral TDF-FTC placebo, (iv) vaginal 1% TFV gel, or (v) vaginal placebo gel. Standard HIV risk-reduction counselling, individualized adherence counselling, condoms, and hepatitis B immunization were provided.<br><br>Regarding contraception use during the study, the protocol laid out that "All participants will complete monthly follow-up visits for a period of 12 – 33 months and will receive ongoing HIV risk reduction counseling, condoms, and diagnosis and treatment of STIs throughout the course of study participation".<br><br>**Outcome:**<br>Prevention of HIV incidence<br><br>**Any significant effect(s):**<br>No. The interventions did not change HIV incidence significantly. The effectiveness was −49.0% with TDF (HR: 1.49 [0.97, 2.29]), −4.4% with TDF-FTC (HR: 1.04 [0.73, 1.49]), and 14.5% with TFV gel (HR: 0.85 [0.61, 1.21]). | | |
| | **HPTN035 [7]**:<br>**Interventions:**<br>4 arms: 0.5% PRO2000 Gel, BufferGel, Placebo Gel, no gel<br><br>Regarding the risk reduction strategies, the study protocol stated that "enrolled participants then complete monthly follow-up visits for the duration of their participation. At each of these visits, participants complete an interval medical and menstrual history and undergo pregnancy testing. HIV/STD risk reduction counseling messages are | | |

| | | | | | | | |
|---|---|---|---|---|---|---|---|
| | reinforced if needed and study supplies (i.e., condoms and the assigned study product, if applicable) are provided".<br><br>**Outcome:**<br>Prevention of HIV incidence<br><br>**Any significant effect(s):**<br>No statistically different effects overall: HIV incidence in the 0.5% PRO2000 gel arm versus the placebo gel arm (HR: 0.7, P = 0.10), and versus the no gel arm (HR: 0.67, P = 0.06); the BufferGel versus placebo gel (HR: 1.10, P = 0.63), and no gel (HR: 1.05, P = 0.78); the placebo gel vs no gel arms (HR: 0.97, P = 0.89). | | | | | | |
| **Balkus (2016) [8]** | **VOICE [6]**<br>**Interventions:**<br>5 arms: (i) oral TDF (300 mg) and TDF-FTC placebo, (ii) oral TDF-FTC (300 mg of TDF and 200 mg of FTC) and TDF placebo, (iii) oral TDF placebo and oral TDF-FTC placebo, (iv) vaginal 1% TFV gel, or (v) vaginal placebo gel. Standard HIV risk-reduction counselling, individualized adherence counselling, condoms, and hepatitis B immunization were provided.<br><br>**Outcome:**<br>Prevention of HIV incidence<br><br>**Any significant effect(s):**<br>No. The interventions did not change HIV incidence significantly. The effectiveness was −49.0% with TDF (HR: 1.49 [0.97, 2.29]), −4.4% with TDF-FTC (HR: 1.04 [0.73, 1.49]), and 14.5% with TFV gel (HR: 0.85 [0.61, 1.21]). | RCT; (i) sexually active women, (ii) willing to use contraception / not planning to get pregnant | Median: 24<br>IQR: 21-29 | **Outcome determination**: Monthly follow-up censored at 1-year<br><br>**Prediction horizon:**<br>1 year | • Age (<25 vs. 25+ yrs)<br>• Married or living with husband or primary partner<br>• Participant earns her own incomes<br>• Number of live births<br>• Alcohol use in the past 3 mo<br>• Partner provides financial or material support<br>• Primary sex partner has other partners<br>• Primary partner is circumcised<br>• Any curable STI<br>• Curable STIs as separate factors<br>• HSV-2 seropositive<br>• participant education level,<br>• primary male partner circumcision status<br>• vaginal sex in the past 4 weeks,<br>• unprotected sex in the past week<br>• number of sex partners in the past 3 months<br>• anal sex in the past 3 months<br>• intravaginal washing with water in the past<br>• intravaginal washing with soap in the past 3 months. | **Only predictors with univariate association with P<0.05 were included.**<br><br>**Variables included:**<br>• Age (<25 vs. 25+ yrs)<br>• Married or living with husband or primary partner<br>• Alcohol use in the past 3 mo<br>• Partner provides financial or material support<br>• Primary sex partner has other partners<br>• Any curable STIs<br>• HSV-2 seropositive<br>• Number of live births | **Cox regression**; Stepwise backward elimination procedures with final model selected based on Akaike information criterion (AIC)<br><br>**Final model:**<br>• Age (<25 vs. 25+ yrs)<br>• Married or living with husband or primary partner<br>• Alcohol use in the past 3 mo<br>• Partner provides financial or material support<br>• Primary sex partner has other partners<br>• Any curable STIs<br>• HSV-2 seropositive |
| **Peebles (2020) [9]** | **ECHO [10]**<br>**Intervention(s):**<br>3 arms: (i) intramuscular depot medroxyprogesterone acetate (DMPA-IM), (ii) a copper intrauterine device (IUD), (iii) a levonorgestrel (LNG) implant<br><br>**Outcome:**<br>HIV incidence<br><br>**Any significant effect(s):**<br>No. HRs for were 1.04 [0.82, 1.33] (p=0.72) for DMPA-IM compared with copper IUD, 1.23 [0.95, 1.59] (p=0.097) for DMPA-IM compared with LNG implant, and 1.18 [0.91, 1.53] (p=0·19) for copper IUD compared with LNG implant<br><br>On HIV risk reduction, site teams consistently counselled participants that none of the three contraceptive methods being used in the study provided protection against HIV or other STIs and advised women to always use condoms in addition to their contraceptive method. | RCT; (i) sexually active women, (ii) seeking effective contraception | Stratified analysis for 25-35 | **Outcome determination**: Follow-up censored at 1-year<br><br>**Prediction horizon:**<br>1 year | • Age (<27 vs. 27+)<br>• Marital/cohabitation status<br>• Weekly alcohol consumption<br>• HIV-1 prevalence (10-15%, 16-20%, 21-25%, 26-30%)<br>• Province (Western Cape, Eastern Cape, KwaZulu-Natal Gauteng, North West)<br>• No. of sex partners in previous 3 months (0 or 1 vs 2+)<br>• Partner has sex with others (No, Yes or do not know)<br>• Condom use (Never or rarely Sometimes, often, or always)<br>• *N. gonorrhoeae*<br>• *C. Trachomatis*<br>• HSV-2 positive<br>• number of previous pregnancies (continuous and categorical),<br>• number of living children (continuous and categorical)<br>• desire for future children<br>• vaginal sex in the past week (Yes/No)<br>• vaginal sex in the past 2 weeks (Yes/No)<br>• number of vaginal sex acts in the past week<br>• vaginal sex during menses in the previous 3 months,<br>• anal sex in the previous 3 months,<br>• partner circumcision status,<br>• partner HIV-1 status,<br>• educational attainment<br>• presence of vaginal discharge<br>• earns own income* (assumed to be considered as one of the candidate predictors since Peebles validated the | **Only predictors with univariate association with P<0.10 were included.**<br><br>**Variables included:**<br>• Age (<27 vs. 27+)<br>• Marital/cohabitation status<br>• Province (Western Cape, Eastern Cape, KwaZulu-Natal Gauteng, North West)<br>• N. gonorrhoeae<br>• C. Trachomatis<br>• HSV-2 positive | **Cox regression**; Stepwise backward elimination procedures with final model selected based on AIC<br><br>**Final model:**<br>• Age (<27 vs. 27+)<br>• Marital/cohabitation status<br>• Province (Western Cape, Eastern Cape, KwaZulu-Natal Gauteng, North West)<br>• *N. gonorrhoeae**<br>• HSV-2 positive*<br><br>* Laboratory-based variables are not included in the modified model |

| | | | | | | | |
|---|---|---|---|---|---|---|---|
| | | | | | VOICE score, and this adds up to 25 predictors claimed by the authors)<br>• Receives material and/or financial support from partner*<br><br>*assumed to be considered as one of the candidate predictors since Peebles validated the VOICE score, and this adds up to 25 predictors claimed by the authors) | | |
| **(II) Adolescent girl and young women (AGYW)** | | | | | | | |
| **Peebles (2020)** | | RCT;<br>(i) female<br>(ii) sexually active<br>(iii) seeking effective contraception | Stratified analysis for 18-24 | **Same as above**. | **Please refer to the list of candidate predictors above.** | • Reported condom use frequency<br>• Marital/cohabitation status<br>• Number of sex partners in the previous 3 months,<br>• Whether a primary partner<br>• has other sex partners<br>• Alcohol consumption<br>• HIV-1 prevalence<br>• N. gonorrhoeae<br>• C. Trachomatis<br>• HSV-2 positive | • Reported condom use frequency<br>• Number of sex partners in the previous 3 months,<br>• Whether a primary partner<br>• has other sex partners<br>• Alcohol consumption<br>• HIV-1 prevalence<br>• N. gonorrhoeae<br>• HSV-2 positive |
| **Burgess (2018) [11]** | **CAPRISA004 [12]**<br>**Intervention(s):**<br>Tenofovir gel versus placebo gel<br><br>**HIV risk reduction:**<br>At enrolment and monthly follow-up visits, participants were provided with comprehensive HIV prevention services (HIV pre- and post-test counselling, HIV risk reduction counselling, condoms, and STI treatment), reproductive health services, and assigned study gel.<br><br>**Outcome:**<br>Prevention of HIV incidence<br><br>**Any effect(s):**<br>Yes. Incidence rate ratio in tenofovir gel versus placebo gel was 0.61 (P = 0.017). | RCT;<br>(i) female<br>(ii) sexually active<br>(iii) agree to use a non-barrier form of contraceptive | Stratified analysis for 18-24 | **Outcome determination**: Anytime within the follow-up period.<br><br>**Time period of prediction**: the entire follow-up period | **Predictors in the original VOICE score:**<br>• Age (<25 vs. 25+ yrs)<br>• Married or living with husband or primary partner<br>• Alcohol use in the past 3 mo<br>• Partner provides financial or material support<br>• Primary sex partner has other partners<br>• Any curable STIs<br>• HSV-2 seropositive<br><br>And other unspecified predictors of interest, including:<br>• Casual partners in last year | No information. | **Cox regression**;<br>AIC was provided for each model<br><br>**Final model:**<br>• Partner(s) has other partners<br>• HSV-2 positive<br>• Casual partners in last year |
| **Rosenberg (2020) [13]** | **Rosenberg [14]**<br>**Intervention(s):**<br>Four service delivery models: (i) standard of care, (ii) Youth-Friendly Health Services (YFHS), (iii) YFHS+behavioural intervention and (iv) YFHS+BI+conditional cash transfer<br><br>**Outcome(s):**<br>HIV and SRH health service utilization<br><br>**Any effect(s):**<br>No effects on HIV incidence were reported. However, 26%, 78%, 80%, and 89% of participants received male or female condoms at least once in models (i)-(iv), respectively. Based on clinical data, 72%, 96%, 100%, and 96% of participants received an HIV test at least once in models (i)-(iv). | Quasi-experimental;<br><br>(i) female<br>(ii) 15 to 24 yrs old,<br>(iii) in the clinic's catchment area,<br>(iv) sexually active | 15-19 yrs old (58.7%)<br><br>20-24 yrs old (41.3%) | **Outcome determination**: Follow-up at 1-year.<br><br>**Prediction horizon:**<br>1 year | **Predictors in the original VOICE score:**<br>• Age (<25 vs. 25+ yrs)<br>• Married or living with husband or primary partner<br>• Alcohol use in the past 3 months<br>• Partner provides financial or material support<br>• Primary sex partner has other partners<br>• Self-reported vaginal discharge (proxy for curable STIs)<br>• Self-reported genital ulcers (proxy for HSV-2 seropositivity)<br><br>**Other candidate predictors**<br>• Being separated, divorced, or widowed<br>• No running water at home<br>• ⩽2 household assets<br>• Being a double orphan<br>• Multiple sexual partners in the last year<br>• Heavy alcohol use<br>• Self-reported vaginal discharge or genital ulcers<br>• Past pregnancy<br>• Partner slept away ⩾3 nights in the last year<br>• Transactional sex<br>• Uncircumcised partner<br>• Perceived partner concurrency<br>• Partner older for > 5 yrs | The authors first performed automatic stepwise backward elimination to determine which VOICE variables to retain using a likelihood ratio test P-value ⩽0.15. They then added in additional candidate predictors with a univariate association of P⩽0.15.<br><br>**Variables included:**<br>• Age<br>• Self-reported genital ulcers<br>• Self-reported vaginal discharge<br>• Primary sex partner has other partners<br>• >5-year partner age difference<br>• Pregnancy history | **Poisson regression;**<br>Model selected using likelihood ratio tests (LRT)<br><br>**Final model:**<br>• Self-reported genital ulcers<br>• Self-reported vaginal discharge<br>• >5-year partner age difference<br>• Pregnancy history |

| | | | | | | | |
|---|---|---|---|---|---|---|---|
| | | | | | • Partner known to be HIV positive | • Divorced, separated, or widowed<br>• ⩾2 sexual partners in the last year<br>• Transactional sex | |
| **(III) General population** | | | | | | | |
| **Kagaayi (2014) [15]** | No intervention; Rakai Community Cohort Study (RCCS) [16] | Cohort;<br><br>(i) 15–49 years old;<br>(ii) sexually active<br>(iii) living in Rakai district | **Mean:**<br>27.0 (F)<br>28.3 (M)<br><br>**SD:**<br>7.8 (F)<br>8.0 (M) | **Outcome determination**: Annual follow-up<br><br>**Prediction horizon:**<br>1 year | • Age<br>• Marital status<br>• Education<br>• Number of sexual partners in the last year<br>• Frequency of condom use<br>• Use of alcohol before sex by either partner<br>• Casual sex<br>• Transactional sex<br>• Concurrent sexual partners<br>• Self-perception of exposure to HIV or perception of exposure by partner<br>• Genital ulcer symptoms<br>• Men's circumcision status<br>• Use of hormonal contraception by women<br>• HIV testing and counselling in the previous 12 months<br>• Community type (trading centre versus village),<br>• Whether one migrated to the community within the previous 2 years<br>• Community HIV prevalence<br>• High risk occupation<br>• Partner in a high-risk occupation<br>• Unknown partner's HIV status | All predictors were included in the initial model | **Cox regression;**<br>Stepwise backward elimination procedures selecting model that minimises the AIC minimisation<br><br>**Final model:**<br><br>**Men**<br>• Age<br>• Education<br>• Circumcision status<br>• Number of sexual partners,<br>• Alcohol consumption by self or partner<br>• Genital ulcers<br>• Being unaware of a partner's HIV status<br>• Community type<br>• Having a partner with a high-risk employment<br>• Community type<br>• Community HIV prevalence<br><br>**Women**<br>• Age,<br>• Marital status,<br>• Education,<br>• Number of sex partners,<br>• Having a new sex partner,<br>• Alcohol consumption by self or partner<br>before sex,<br>• Having concurrent relationships,<br>• Being employed in a high-risk occupation,<br>• Having genital ulcers,<br>• Community HIV prevalence,<br>• Perceiving oneself or partner to have been exposed to HIV infection |
| **Balzer (2020) [17]** | **Sustainable East Africa Research in Community Health (SEARCH) [18]**:<br>**Intervention(s):**<br>Universal antiretroviral therapy (ART) with annual population testing and a multi-disease, patient-centred strategy. Only the intervention arm was included in this analysis.<br><br>**Outcome(s):**<br>Prevention of HIV incidence and improvement of community health<br><br>**Any effect(s):** No significant effect on HIV incidence (relative risk: 0.95 [0.77, 1.17]) | RCT;<br>(i) 15+ years old<br>(ii) community residents | 15–24 (39%)<br><br>25–34 (20%)<br><br>35–44 (15%)<br><br>45–54 (11%)<br><br>55+ (16%) | **Outcome determination**: Annual follow-up<br><br>**Prediction horizon:**<br>1 year | **DEMOGRAPHY**<br>• Age<br>• Sex<br>• Marital status<br>• Polygamy<br>• Familial relation to head-of-household<br>• Education<br>• Occupation strata<br>• Student<br>• Transportation<br>• Fisherman or fishmonger<br>• Bar worker<br>• Hotel worker<br>• Shopkeeper<br>• Alcohol use<br>• Region | **Method (1) "Risk group approach":**<br>• women aged 15–24 years<br>• individuals with spouses who were living with HIV<br>• alcohol users<br>• widow(er)s<br>• persons employed in transportation<br>• bar worker<br>• fisherman<br><br>**Method (2) "Model-based approach":**<br>All candidate predictors with univariable | **Method (1) "Risk group approach":** the risk score is computed as the sum of the groups (list on the left) to which an individual belongs.<br><br>**Method (2) "Model-based approach":** a logistic model with forward and backward stepwise selection.<br><br>**Method (3) "Machine-learning approach":** Super-learner ensemble method [17] |

| | | | |
|---|---|---|---|
| | | **MOBILITY**<br>• Immigrant<br>• Baseline stable resident<br>• Mobile resident<br>• Shifted residence<br>• Nights away<br>**HEALTH**<br>• Health fair attendance<br>• Contraceptive use<br>• Pregnant<br>• Live birth<br>• Male circumcision<br><br>**SPOUSES**<br>• Unknown status<br>• Serodiscordant<br>• Serodiscordant and male (Spouse is HIV-infected and male)<br>• Serodiscordant and circumcision (Spouse is HIV-infected and male partner is not circumcised)<br>• Serodiscordant and polygamous (Spouse is HIV-infected and the marriage is polygamous)<br>• Serodiscordant and unsuppressed (Spouse is HIV-infected with HIV RNA level >500 copies/mL)<br><br>**HOUSEHOLD FACTORS**<br>• Wealth<br>• HIV-unknown adult (At least 1 adult whose HIV status is unknown in the household)<br>• HIV-infected adult (At least 1 HIV-infected adult in the household)<br>• HIV-infected adult of the opposite sex (At least 1 adult of the opposite sex and HIV-infected in the household)<br><br>**INTERACTIONS**<br>• Young woman (Woman aged 15-24 years)<br>• Female bar worker<br>• Wealthy male<br>• Young pregnancy (Woman aged 15-24 years and reporting current pregnancy)<br>• Young mother (Woman aged 15-24 years and reporting at least 1 live birth in the past year) | association of P<0.05 were included<br><br>**Method (3) “Machine-learning approach”:** All candidate predictors |

| | | | | | |
|---|---|---|---|---|---|
| **Roberts (2021) [19]** | No intervention. Africa Centre Demographic Information System (ACDIS) cohort [20] | Cohort;<br>(i) Resident members aged 15+ years old | **INDIVIDUAL-LEVEL**<br>• age<br>• sex<br>• marital status<br>• employment<br>• education<br>• socioeconomic status<br>• migration history<br>• ever had sex<br>• prior pregnancy/children (women)<br>• contraception use (women)<br>• circumcision status (men)<br>• number of partners in last 12 months<br>• current number of partners<br>• most recent partner's age<br>• used condom at last sex<br>• most recent partner type (causal vs. regular)<br>• most recent partner residence (same household vs. outside of household)<br>**GEOSPATIAL COVARIATES**<br>• local HIV prevalence (by year)<br>• local population prevalence of detectable viremia (by year)<br>• distance to roads<br>• distance to clinics<br>• distance to schools<br>• urban/rural. | All candidate predictors were included | Cox proportional hazards regression models with lasso penalties and time-varying covariates. Penalties selected via cross-validation. |

#### Table S2. Methods for assessing curable sexually-transmitted infections (STIs) and HSV-2 status.

| First author (Year) | Score | Dev/ Val | Sources of data | Variable included in the model | Description | Methods of assessment | Enrolment | Ref |
|---|---|---|---|---|---|---|---|---|
| **Wand (2012) [1]** | Self-derived | Dev | MIRA | Genital epithelial disruption | Presence of clinically apparent lesion(s) with epithelial disruption at the baseline | - | - | [21] |
| | | | | Genital discharge | Presence of genital discharge at the baseline | - | - | - |
| **Wand (2018) [3]** | Self-derived | Dev | MIRA | Diagnosed with STIs (at least one of chlamydia, gonorrhoea, or syphilis) at the baseline | Laboratory tests at enrolment | • PCR for chlamydia<br>• PCR for gonorrhoea<br>• PCR for Trichomonas | • If positive, participants needed to complete treatment before enrolment | [2] |
| | | | MDP301 | | Laboratory tests at enrolment | • PCR for chlamydia<br>• PCR for gonorrhoea<br>• Laboratory test for trichomonas | • At enrolment, women who had clinically suspicious cervical lesion were referred for assessment and excluded from the study | [4, 22] |
| | | | NCT00213083 | | Laboratory tests at enrolment | • Laboratory test for chlamydia, gonorrhoea, syphilis | - | [5] |
| | | | VOICE | | Laboratory tests at enrolment | • Urine strand displacement amplification (SDA) for chlamydia<br>• Urine SDA for gonorrhoea<br>• Serology test for syphilis<br>• Rapid test for trichomonas | • At enrolment, women tested positive for STIs were offered treatment and might be enrolled after treatment is completed and symptoms have resolved within 56 days of obtaining informed consent. | [6] |
| | | | HPTN 035 | | Laboratory tests at enrolment | • Urine SDA for chlamydia<br>• Urine SDA for gonorrhoea<br>• Serology test for syphilis<br>Wet mount for trichomonas | • Women tested positive for STIs were not enrolled unless treatment was completed, and all symptoms had resolved within 30 days of obtaining informed consent for screening. | [7] |
| **Balkus (2016) [8]** | VOICE | Dev | VOICE | STIs at baseline | Laboratory tests at enrolment | • Urine SDA for chlamydia<br>• Urine SDA for gonorrhoea<br>• Serology test for syphilis<br>• Rapid test for trichomonas | • At enrolment, women tested positive for STIs were offered treatment and might be enrolled after treatment is completed and symptoms have resolved within 56 days of obtaining informed consent. | [6] |
| | | | | HSV-2 | Laboratory tests at enrolment | • Serology test for HSV-2 | | [6] |
| | | Val | HPTN 035 | STIs at baseline | Laboratory tests at enrolment | • Urine SDA for chlamydia<br>• Urine SDA for gonorrhoea<br>• Serology test for syphilis<br>• Wet mount for trichomonas | • Women tested positive for STIs were not enrolled unless treatment was completed, and all symptoms had resolved within 30 days of obtaining informed consent for screening. | [23] |
| | | | | HSV-2 | Laboratory tests at enrolment | • Serology test for HSV-2 | | [23] |
| | | Val | FEM-PrEP | STIs at baseline | Laboratory tests at enrolment | • PCR for chlamydia<br>• PCR for gonorrhoea<br>• Serology test for syphilis<br>• Wet mount for trichomonas | | [24] |
| | | | | HSV-2 | Laboratory tests at enrolment | • Serology test for HSV-2 | | [24] |

| | | | | | | | |
|---|---|---|---|---|---|---|---|
| **Balkus (2018) [25]** | VOICE | Val | ASPIRE | STIs at baseline | Laboratory tests | • PCR for chlamydia<br>• PCR for gonorrhoea<br>• Serology test for syphilis<br>• Rapid test for trichomonas | [26] |
| | | | | No HSV-2 collected | - | - | - |
| **Burgess (2017) [27]** | VOICE | Val | FACTS 001 | Self-reported STIs | Syndromic management assessed by study staff` | - | |
| | | | | HSV-2 | Laboratory tests | • HSV-2 status established at enrolment according to a testing algorithm in the protocol. Incident cases were confirmed by HSV Western blot. | [28] |
| **Burgess (2018) [11]** | VOICE + self | Val + Dev | CAPRISA 004 | STIs at baseline | Self-report and syndromic management | - | |
| | | | | HSV-2 | Laboratory tests | • Serology test (IgG ELISA) for HSV-2 | [29] |
| **Peebles (2020) [9]** | VOICE + self | Val + Dev | ECHO | STIs at baseline | Laboratory tests | • PCR for chlamydia<br>• PCR for gonorrhoea | [30] |
| | | | | HSV-2 | Laboratory tests | • Serology test for HSV-2 | |
| **Giovenco (2019) [31]** | VOICE | Val | HPTN 068 | No curable STIs collected | - | - | |
| | | | | HSV-2 | Laboratory tests | • Serology test (IgG ELISA) for HSV-2 | [32] |
| **Rosenberg (2020) [13]** | VOICE + self | Val + Dev | Girl Power - Malawi | STIs at baseline | Self-reported abnormal vaginal discharge in the last 6 months | - | |
| | | | | HSV-2 | Self-reported genital sores in the last 6 months | -. | |
| **Ayton (2020) [33]** | VOICE + self | Val | CAPRISA 007 | No curable STIs collected | - | - | |
| | | | | HSV-2 | Laboratory tests | • Serology test (IgG ELISA) for HSV-2 measured at baseline, 12 months and 24 months. | [34] |
| **Kagaayi (2014) [35]** | - | Dev | RCCS | Genital ulcer symptoms | Self-reported genital ulcer symptoms in the past 12 months | - | |
| **Balzer (2019) [17]** | - | Dev | SEARCH | No STIs collected | - | - | |
| **Roberts (2021) [19]** | - | Dev | ACDIS | No STIs collected | - | - | |

#### Table S3. Summary table for the risk of bias assessment according to the PROBAST checklist

| First author (Year) | Wand (2012) | Wand (2018) | Balkus (2016) | Peebles (2020) 18-24 | Peebles (2020) 25-35 | Burgess (2018) | Rosenberg (2020) | Kagaayi (2014) Men | Kagaayi (2014) Women | Balzer (2020) | Roberts (2021) Men | Roberts (2021) Women | Wand (2018) | Balkus (2016) | Balkus (2018) | Peebles (2020) | Burgess (2017) | Burgess (2018) | Giovenco (2019) | Ayton (2020) | Rosenberg (2020) |
|---|---|---|---|---|---|---|---|---|---|---|---|---|---|---|---|---|---|---|---|---|---|
| | Article | Article | Article | Article | | Poster | Article | Article | | Article | Abstract | | Article | Article | Letter | Article | Poster | Poster | Article | Article | Art |
| | Development | | | | | | | | | | | | Validation | | | | | | | | |
| Participants | | | | | | | | | | | | | | | | | | | | | |
| 1.1 | Y | Y | Y | Y | Y | Y | Y | Y | Y | Y | Y | Y | Y | Y | Y | Y | Y | Y | Y | Y | Y |
| 1.2 | Y | Y | Y | Y | Y | Y | PY | Y | Y | Y | Y | Y | Y | Y | Y | Y | Y | Y | Y | Y | PY |
| Overall | Low | Low | Low | Low | Low | Low | Low | Low | Low | Low | Low | Low | Low | Low | Low | Low | Low | Low | Low | Low | Low |
| Predictors | | | | | | | | | | | | | | | | | | | | | |
| 2.1 | Y | Y | Y | Y | Y | Y | Y | Y | Y | Y | Y | Y | Y | Y | Y | Y | Y | Y | Y | Y | Y |
| 2.2 | Y | Y | Y | Y | Y | Y | Y | Y | Y | Y | Y | Y | Y | Y | Y | Y | Y | Y | Y | Y | Y |
| 2.3 | Y | Y | Y | Y | Y | Y | Y | Y | Y | Y | Y | Y | N | N | N | Y | PY | Y | N | N | PY |
| Overall | Low | Low | Low | Low | Low | Low | Low | Low | Low | Low | Low | Low | High | High | High | Low | Low | Low | High | High | Low |
| Outcome | | | | | | | | | | | | | | | | | | | | | |
| 3.1 | Y | Y | Y | Y | Y | Y | Y | Y | Y | Y | Y | Y | Y | Y | Y | Y | Y | Y | Y | Y | Y |
| 3.2 | Y | Y | Y | Y | Y | Y | Y | Y | Y | Y | Y | Y | Y | Y | Y | Y | Y | Y | Y | Y | Y |
| 3.3 | Y | Y | Y | Y | Y | Y | Y | Y | Y | Y | Y | Y | Y | Y | Y | Y | Y | Y | Y | Y | Y |
| 3.4 | Y | Y | Y | Y | Y | Y | N | Y | Y | Y | Y | Y | Y | Y | Y | Y | Y | Y | Y | Y | N |
| 3.5 | PY | PY | PY | PY | PY | PY | PY | PY | PY | PY | PY | PY | PY | PY | PY | PY | PY | PY | PY | PY | PY |
| 3.6 | Y | PN | Y | Y | Y | PY | Y | Y | Y | N | Y | Y | NI | Y | Y | Y | PN | PY | Y | Y | Y |
| Overall | Low | Low | Low | Low | Low | Low | High | Low | Low | High | Low | Low | Unclear | Low | Low | Low | Low | Low | Low | Low | High |
| Analysis | | | | | | | | | | | | | | | | | | | | | |
| 4.1 | N | Y | Y | N | N | NI | N | Y | Y | Y | Y | Y | NI | Y | Y | Y | PY | N | N | N | N |
| 4.2 | N | N | Y | Y | N | Y | N | Y | Y | Y | NI | NI | NI | Y | Y | Y | Y | Y | Y | PY | Y |
| 4.3 | NI | NI | Y | Y | Y | Y | N | NI | NI | NI | NI | NI | NI | Y | Y | Y | N | Y | N | N | N |
| 4.4 | NI | NI | N | Y | Y | Y | N | Y | Y | N | Y | Y | NI | N | Y | Y | N | Y | N | N | N |
| 4.5 | N | N | N | N | N | NI | N | Y | Y | N | Y | Y | | | | | | | | | |
| 4.6 | Y | Y | Y | Y | Y | Y | PY | Y | Y | N | Y | Y | NI | Y | Y | Y | Y | Y | Y | Y | Y |
| 4.7 | NI | N | NI | Y | Y | NI | NI | Y | Y | NI | Y | Y | NI | Y | Y | Y | Y | Y | Y | PY | Y |
| 4.8 | N | Y | N | Y | Y | N | N | N | N | N | Y | Y | | | | | | | | | |
| 4.9 | Y | Y | Y | Y | Y | Y | Y | Y | Y | NI | NI | NI | | | | | | | | | |
| Overall | High | High | High | Low | High | Unclear | High | Unclear | Unclear | Unclear | Unclear | Unclear | Unclear | High | Low | Low | High | Low | High | High | High |
| Overall | High | High | High | Low | High | Unclear | High | Unclear | Unclear | Unclear | Unclear | Unclear | Unclear | High | High | Low | High | Low | High | High | High |

To identify information beyond what were given by the risk score articles, we referred to the original sources of data, including the clinical trial protocols, and the methodological articles.

#### Table S4. Summary table on the concerns for applicability according to the PROBAST checklist

| Wand (2012) | Wand (2018) | Balkus (2016) | Burgess (2018) | Peebles (2020) 18-24 | Peebles (2020) 25-35 | Rosenberg (2020) | Kagaayi (2014) Men | Kagaayi (2014) Women | Balzer (2019) | Roberts (2021) Men | Roberts (2021) Women | Wand (2018) | Balkus (2016) | Balkus (2018) | Burgess (2017) | Burgess (2018) | Peebles (2020) | Giovenco (2019) | Ayton (2020) | Rosenberg (2020) |
|---|---|---|---|---|---|---|---|---|---|---|---|---|---|---|---|---|---|---|---|---|
| Development | | | | | | | | | | | | Validation | | | | | | | | |
| (I) Participants | | | | | | | | | | | | | | | | | | | | |
| Low | Low | Low | Low | Low | Low | Low | Low | Low | Low | Low | Low | Low | Low | Low | Low | Low | Low | Low | Low | Low |
| (II) Predictors | | | | | | | | | | | | | | | | | | | | |
| Low | High | High (incl STIs)<br>Low (excl STIs) | High (incl HSVs)<br>Low (excl HSVs) | High (incl STIs)<br>Low (excl STIs) | High (incl STIs)<br>Low (excl STIs) | Low | Low | Low | High | Low | Low | Low | High (incl STIs)<br>Low (excl STIs) | High | High | High | High (incl STIs) | High | High | Low |
| (III) Outcome | | | | | | | | | | | | | | | | | | | | |
| Low | High | Low | Low | Low | Low | Low | Low | Low | Low | Low | Low | Uncertain | Low | Low | High | Low | Low | Low | High | Low |
| Overall | | | | | | | | | | | | | | | | | | | | |
| Low | High | High (full)<br>Low (excl STIs) | High (incl STIs)<br>Low (excl STIs) | High (incl STIs)<br>Low (excl STIs) | High (incl STIs)<br>Low (excl STIs) | High | Low | Low | Low | Low | Low | Uncertain | High (incl STIs)<br>Low (excl STIs) | High | High | High | High (incl STIs) | High | High | High |

#### Table S5. Summary of missing data and loss-to-follow-up

| Author, Year | Ayton 2020 | Burgess 2017 | Burgess 2018 | Balkus 2016 | | | Balkus 2018 | Balzer 2020 | Kagaayi 2014 | Giovenco 2019 | Peebles 2020 | Roberts 2021 | Rosenberg 2020 | Wand 2012 | Wand 2018 |
|---|---|---|---|---|---|---|---|---|---|---|---|---|---|---|---|
| Cohort name | CAPRISA 007 | FACTS 001 | CAPRISA 004 | VOICE | HPTN035 | FEM-PrEP | ASPIRE | SEARCH | RCCS | HPTN068 | ECHO | ACDIS | Girl Power | MIRA | Multiple |
| (I) Enrollment | | | | | | | | | | | | | | | |
| Total enrolled (excl ineligible if information available) | 1049 | NI | NI | 5007 | 3101 | 2120 | 2629 | NI | 21957 | 2533 | 5670 | NI | 1000 | NI | NI |
| (II) Analysis | | | | | | | | | | | | | | | |
| Imcomplete data | 78 | 944 | 13 | 38 | NI | NI | NI | NI | NI | 355 | 99 | NI | 172 | NI | NI |
| Loss to follow up | 40 | NI | NI | 142 | NI | NI | 90 | NI | 8677 | 30 | 97 | NI | NI | NI | NI |
| Included in the final analysis | 971 | 1115 | 431 | 4834 | 2848 | 1804 | 2539 | 75558 | 13280 | 2178 | 5573 | 19556 | 795 | 1485 | 8982 |
| % analysed | 92.6% | NI | NI | 96.5% | 91.8% | 85.1% | 96.6% | NI | 60.5% | 86.0% | 98.3% | NI | 79.5% | NI | NI |
| (III) Handling missing data | | | | | | | | | | | | | | | |
| Missing predictors | Imputed | All predictors collected | All predictors collected | N/A (Develop-ment study) | NI | NI | NI | N/A (Develop-ment study) | N/A (Develop-ment study) | NI | All predictors collected | N/A (Develop-ment study) | All predictors collected | N/A (Develop-ment study) | N/A (Develop-ment study) |
| Missing baseline or outcome data | NI | NI | NI | Missing <5% | NI | NI | NI | Imputed | Imputed | NI | Missing <5% | Imputed | NI | NI | NI |

Abbreviation: (NI) No information

#### Table S6. Summary adjusted and unadjusted hazard ratios (HRs)

| | | Women; sexually active and seeking effective contraception (RCTs) | | | | | | AGYW only | | | | | |
|---|---|---|---|---|---|---|---|---|---|---|---|---|---|
| | | No. of studies | Pooled estimates | 95% CI | Between-studies var | $I^2$ | Ref | No. of studies | Pooled estimates# | 95% CI | Between-studies var | $I^2$ | Ref‡ |
| **Adjusted effects ¶** | Younger age§ | 5 | 1.62 | [1.17, 2.23] | 0.0924 | 53.1% | [3, 8, 9, 11, 27] | - | - | - | - | - | - |
| | Not married /cohabiting | 6 | 2.33 | [1.73, 3.13] | 0.0553 | 41.3% | [1, 3, 8, 9, 11, 27] | 1 | 1.57# | [0.89;3.09] | - | - | [9] |
| | No. of sex partners | 2 | 1.62 | [1.27; 2.07] | 0.0061 | 0.0% | [1, 3] | 2 | 1.76 | [1.19; 2.60] | 0.0102 | 0.0% | [9, 11] |
| | Partners having other partners | 3 | 1.67 | [1.04; 2.71] | 0.2322 | 51.3% | [8, 27] | 2 | 2.35 | [0.48; 11.53] | 0.9476 | 65.3% | [9, 11] |
| | Curable STIs | 6 | 1.45 | [1.17; 1.79] | 0.0290 | 0.0% | [1, 3, 8, 9, 11, 27] | 2 | 2.14 | [1.40; 3.25] | 0.0018 | 0.0% | 11,13 |
| | HSV-2 | 4 | 1.67 | [1.34; 2.09] | 0.0076 | 0.0% | [8, 9, 11, 27] | 3 | 1.77 | [1.24; 2.54] | 0.0299 | 16.1% | [9, 11, 13] |
| **Unadjusted effects§** | Younger age | 3 | 1.71 | [1.31, 2.22] | 0.0180 | 0.0% | [1, 8, 11] | - | - | - | - | - | - |
| | Not married /cohabiting | 4 | 1.90 | [1.25, 2.87] | 0.0979 | 36.7% | [1, 8, 11, 27] | 2 | 0.76 | [0.21, 2.75] | 0.3974 | 43.5% | [13, 31] |
| | No. of sex partners | 2 | 2.02 | [1.44; 2.82] | 0.0023 | 0.0% | [8, 27] | 1 | 2.34 | [0.78; 7.00] | - | - | 13 |
| | Partners having other partners | 2 | 2.18 | [1.51; 3.13] | 0.0414 | 0.0% | [8, 27] | 2 | 1.83 | [0.97; 3.48] | 0.0048 | 0.0% | 13,31 |
| | Curable STIs | 3 | 1.52 | [1.04, 2.21] | 0.0663 | 50.7% | [1, 8, 11] | 1 | 3.36# | [1.17, 9.68] | - | - | [13] |
| | HSV-2 | 3 | 1.47 | [1.22; 2.59] | 0.0002 | 0.0% | [8, 11, 27] | 2 | 2.85 | [1.31; 6.22] | 0.0001 | 0.0% | 13,31 |

¶ Adjusted HR estimates were only available for predictors retained in the final model after model selection.

### For risk factor estimates that were provided by less than two studies, estimates from the single studies were reported.

§ For all age women studies, all used the <25 cut-off, except for Peebles et al., where they used <27 cut-off for the >25 yrs old.

† Fewer studies provided estimates for unadjusted HR than adjusted HR (listed above). For general women studies, Wand (2018)[3], and Peebles[9], did not provide unadjusted HRs; for AGYW studies, Peebles[9], did not provide unadjusted HRs, while Burgess (2018)[11], provided unadjusted estimates for some factors but not all.

‡ Unlike other studies, Rosenberg[13] used incidence rate ratio estimates rather than hazard ratio.

**Table S7. HIV incidence and distribution of high-risk group by each risk score**

| Score name | Dev/ val | First author (Year) | Sources of data | Highest score available | Score | Observed incidence (per 100 py) | Study population | | Incident cases | |
|---|---|---|---|---|---|---|---|---|---|---|
| | | | | | | | % within each score | Cumulative % of high-risk population | % within each score | Cumulative % of incident cases |
| **(I) Women only (All ages / 25+years old)** | | | | | | | | | | |
| VOICE | Dev | Balkus (2016) | VOICE | 11 | 9+ | 14.7 | 8.0% | 8.0% | 18.6% | 18.6% |
| | | | | | 8 | 11.9 | 9.2% | 17.2% | 17.5% | 36.1% |
| | | | | | 7 | 10.4 | 16.3% | 33.4% | 27.4% | 63.5% |
| | | | | | 6 | 5.5 | 18.9% | 52.3% | 17.1% | 80.6% |
| | | | | | 5 | 5.4 | 11.3% | 63.6% | 10.3% | 90.9% |
| | | | | | 4 | 1.9 | 18.2% | 81.8% | 5.7% | 96.6% |
| | | | | | 3 | 1.7 | 4.1% | 85.9% | 1.1% | 97.7% |
| | | | | | 2 | 0.8 | 11.0% | 96.9% | 1.5% | 99.2% |
| | | | | | 1 | 4.1 | 0.6% | 97.4% | 0.4% | 99.6% |
| | | | | | 0 | 0.9 | 2.6% | 100.0% | 0.4% | 100.0% |
| VOICE | Val | Balkus (2016) | HPTN035 | 10 | 8+ | 8.8 | 6.0% | 6.0% | 15.3% | 15.3% |
| | | | | | 7 | 7.8 | 7.2% | 13.2% | 16.3% | 31.6% |
| | | | | | 6 | 6.0 | 10.4% | 23.7% | 18.4% | 50.0% |
| | | | | | 5 | 4.9 | 5.7% | 29.3% | 8.2% | 58.2% |
| | | | | | 4 | 3.1 | 21.2% | 50.6% | 19.4% | 77.6% |
| | | | | | 3 | 4.8 | 4.3% | 54.9% | 6.1% | 83.7% |
| | | | | | 2 | 1.4 | 29.1% | 84.0% | 12.2% | 95.9% |
| | | | | | 1 | 0.0 | 1.3% | 85.3% | 0.0% | 95.9% |
| | | | | | 0 | 0.9 | 14.7% | 100.0% | 4.1% | 100.0% |
| VOICE | Val | Balkus (2016) | FEM-PrEP | 4 | 4 | 7.3 | 8.5% | 8.5% | 11.9% | 11.9% |
| | | | | | 3 | 6.5 | 28.1% | 36.6% | 35.6% | 47.5% |
| | | | | | 2 | 5.4 | 32.5% | 69.1% | 35.6% | 83.1% |
| | | | | | 1 | 2.0 | 25.2% | 94.2% | 11.9% | 94.9% |
| | | | | | 0 | 3.8 | 5.8% | 100.0% | 5.1% | 100.0% |
| VOICE | Val | Balkus (2018) | ASPIRE | 8 | 8 | 10.7 | 1.5% | 1.5% | 4.2% | 4.2% |
| | | | | | 7 | 10.0 | 8.0% | 9.6% | 21.1% | 25.3% |
| | | | | | 6 | 5.7 | 15.9% | 25.5% | 24.2% | 49.5% |
| | | | | | 5 | 3.2 | 9.7% | 35.1% | 8.4% | 57.9% |
| | | | | | 4 | 4.0 | 23.7% | 58.8% | 25.3% | 83.2% |
| | | | | | 3 | 4.6 | 6.0% | 64.8% | 7.4% | 90.5% |
| | | | | | 2 | 1.3 | 27.8% | 92.6% | 9.5% | 100.0% |
| | | | | | 1 | 0.0 | 1.1% | 93.7% | 0.0% | 100.0% |
| | | | | | 0 | 0.0 | 6.3% | 100.0% | 0.0% | 100.0% |

| Score name | | First author (Year) | Sources of data | Highest score available | Score | Observed incidence (per 100 py) | Study population | | Incident cases | |
|---|---|---|---|---|---|---|---|---|---|---|
| | | | | | | | % within each score | Cumulative % of high-risk population | % within each score | Cumulative % of incident cases |
| VOICE | Val | Burgess* (2017) | FACTS001 | 11 | 9+ | 6.1 | 10.1% | 10.1% | 13.6% | 13.6% |
| | | | | | 8 | 4.3 | 11.0% | 21.2% | 11.1% | 24.7% |
| | | | | | 7 | 4.9 | 20.7% | 41.9% | 23.5% | 48.2% |
| | | | | | 6 | 5.6 | 12.4% | 54.3% | 16.1% | 64.2% |
| | | | | | 5 | 3.6 | 23.2% | 77.5% | 19.8% | 84.0% |
| | | | | | 4 | 4.7 | 8.1% | 85.6% | 8.6% | 92.6% |
| | | | | | 3 | 2.0 | 8.0% | 93.5% | 3.7% | 96.3% |
| | | | | | 2 | 2.5 | 4.5% | 98.0% | 2.5% | 98.8% |
| | | | | | 1 | 3.6 | 1.4% | 99.4% | 1.2% | 100.0% |
| | | | | | 0 | 0.0 | 0.6% | 100.0% | 0.0% | 100.0% |
| VOICE | Val | Burgess¶* (2018) | CAPRISA 004 | 11 | 11 | 17.0 | 0.9% | 0.9% | - | - |
| | | | | | 10+ | >30 | 7.7% | 8.6% | - | - |
| | | | | | 9+ | 13.0 | 15.3% | 23.9% | - | - |
| | | | | | 8+ | 7.5 | 22.0% | 45.9% | - | - |
| | | | | | 7+ | 6.0 | 29.2% | 75.2% | - | - |
| | | | | | 6+ | 5.2 | 8.6% | 83.8% | - | - |
| | | | | | 5+ | 5.2 | 11.6% | 95.4% | - | - |
| | | | | | 4+ | 5.5 | 2.6% | 97.9% | - | - |
| | | | | | 3+ | 8.0 | 1.9% | 99.8% | - | - |
| | | | | | 2+ | 0.0 | 0.2% | 100.0% | - | - |
| | | | | | 1+ | 0.0 | 0.0% | 100.0% | - | - |
| | | | | | 0+ | 0.0 | 0.0% | 100.0% | - | - |
| Self-derived | Dev | Peebles¶ (2020) | ECHO | 7 | 7 | 12.0 | 0.8% | 0.8% | 2.8% | 2.8% |
| | | | | | 6 | 8.5 | 16.1% | 16.9% | 39.9% | 42.7% |
| | | | | | 5 | 2.9 | 41.2% | 58.1% | 35.9% | 78.6% |
| | | | | | 4 | 2.3 | 28.9% | 87.0% | 18.6% | 97.2% |
| | | | | | 3 | 0.5 | 8.8% | 95.8% | 1.4% | 98.6% |
| | | | | | 2 | 1.1 | 4.2% | 100.0% | 1.4% | 100.0% |
| | | | | | 1 | 0.0 | 0.0% | 100.0% | 0.0% | 100.0% |

| Score name | Dev/ val | First author (Year) | Sources of data | Highest score possible | Score | Observed incidence (per 100 py) | Study population | | Incident cases | |
|---|---|---|---|---|---|---|---|---|---|---|
| | | | | | | | % within each score | Cumulative % of high-risk population | % within each score | Cumulative % of incident cases |
| **(II) AGYW** | | | | | | | | | | |
| VOICE | Val | Giovenco (2019) | HPTN 068 | 10 | 10 | 0.0 | 0.1% | 0.1% | 6.1% | 6.1% |
| | | | | | 9 | 0.0 | 0.5% | 0.6% | 0.0% | 6.1% |
| | | | | | 8 | 7.1 | 1.2% | 1.7% | 9.1% | 15.2% |
| | | | | | 7 | 1.9 | 6.2% | 7.9% | 45.5% | 60.6% |
| | | | | | 6 | 2.2 | 14.7% | 22.7% | 24.2% | 84.9% |
| | | | | | 5 | 0.9 | 71.2% | 93.9% | 9.1% | 93.9% |
| | | | | | 4 | 2.3 | 5.4% | 99.3% | 6.1% | 100.0% |
| | | | | | 3 | 0.0 | 0.1% | 99.4% | 0.0% | 100.0% |
| | | | | | 2 | 12.9 | 0.6% | 100.0% | 0.0% | 100.0% |
| VOICE | Val | Ayton (2020) | CAPRISA 007 | 6 | 6 | 0.0 | 0.1% | 0.1% | 0.0% | 0.0% |
| | | | | | 5 | 8.0 | 2.6% | 2.7% | 14.3% | 14.3% |
| | | | | | 4 | 1.4 | 7.2% | 9.9% | 7.1% | 21.4% |
| | | | | | 3 | 1.0 | 21.2% | 31.1% | 14.3% | 35.7% |
| | | | | | 2 | 1.4 | 68.8% | 99.9% | 64.3% | 100.0% |
| | | | | | 1 | 0.0 | 0.0% | 99.9% | 0.0% | 100.0% |
| | | | | | 0 | 0.0 | 0.1% | 100.0% | 0.0% | 100.0% |
| Self-derived | Dev | Burgess[¶]* (2018) | CAPRISA 004 | 9 | 9 | 16.5 | 1.4% | 1.4% | - | - |
| | | | | | 8 | >30 | 11.3% | 12.7% | - | - |
| | | | | | 7 | 13.0 | 19.2% | 32.0% | - | - |
| | | | | | 6 | 11.0 | 24.1% | 56.0% | - | - |
| | | | | | 5 | 5.3 | 33.0% | 89.0% | - | - |
| | | | | | 4 | 8.5 | 5.5% | 94.5% | - | - |
| | | | | | 3 | 0.0 | 5.2% | 99.7% | - | - |
| | | | | | 2 | 0.0 | 0.3% | 100.0% | - | - |
| | | | | | 1 | 0.0 | 0.0% | 100.0% | - | - |
| | | | | | 0 | 0.0 | 0.0% | 100.0% | - | - |

| Score name | Dev/ Val | First author (Year) | Sources of data | Highest score possible | Score | Observed incidence (per 100 py) | Study population | | Incident cases | |
|---|---|---|---|---|---|---|---|---|---|---|
| | | | | | | | % within each score | Cumulative % of high-risk population | % within each score | Cumulative % of incident cases |
| Self-derived | Dev | Peebles¶ (2020) | ECHO | 11 | 11 | 0 | 0.0% | 0.0% | 0.0% | 0.0% |
| | | | | | 10 | 28.6 | 0.3% | 0.3% | 0.7% | 0.7% |
| | | | | | 9 | 15.5 | 0.6% | 0.9% | 1.7% | 2.4% |
| | | | | | 8 | 12.4 | 1.9% | 2.8% | 4.3% | 6.7% |
| | | | | | 7 | 12.5 | 3.1% | 5.9% | 7.1% | 13.8% |
| | | | | | 6 | 9.2 | 5.9% | 11.8% | 9.8% | 23.6% |
| | | | | | 5 | 7.2 | 18.7% | 30.5% | 25.0% | 48.6% |
| | | | | | 4 | 4.9 | 33.8% | 64.3% | 31.2% | 79.8% |
| | | | | | 3 | 3.6 | 20.4% | 84.7% | 14.3% | 94.1% |
| | | | | | 2 | 2.5 | 10.7% | 95.4% | 4.8% | 98.9% |
| | | | | | 1 | 1.3 | 4.6% | 100.0% | 1.1% | 100.0% |
| VOICE | Val | Rosenberg¶* (2020) | Girls Power | 11 | 9+ | 5.3 | - | - | - | - |
| | | | | | 8 | 1.5 | - | - | - | - |
| | | | | | 7 | 1.3 | - | - | - | - |
| | | | | | 6 | 5.5 | - | - | - | - |
| | | | | | 5 | 1.0 | - | - | - | - |
| | | | | | 4 | 0.5 | - | - | - | - |
| Self-derived | Dev | | | 5 | 5 | 22.8 | - | - | - | - |
| | | | | | 4 | 6.0 | - | - | - | - |
| | | | | | 3 | 4.5 | - | - | - | - |
| | | | | | 2 | 1.8 | - | - | - | - |
| | | | | | 1 | 1.3 | - | - | - | - |
| **(III) General population** | | | | | | | | | | |
| Self-derived (men) | Dev | Kagaayi (2017) | RCCS (men) | 280 | 179+ | - | 20.0% | 20.0% | 46.7% | 46.7% |
| | | | | | 175+ | - | 5.0% | 25.0% | 8.4% | 55.1% |
| | | | | | 169+ | - | 8.3% | 33.3% | 8.0% | 63.1% |
| | | | | | 164+ | - | 6.7% | 40.0% | 7.1% | 70.2% |
| | | | | | 158+ | - | 10.0% | 50.0% | 8.5% | 78.7% |
| Self-derived (women) | Dev | Kagaayi (2017) | RCCS (women) | 500 | 325+ | - | 20.0% | 20.0% | 40.1% | 40.1% |
| | | | | | 314+ | - | 5.0% | 25.0% | 7.9% | 48.0% |
| | | | | | 298+ | - | 8.3% | 33.3% | 7.6% | 55.6% |
| | | | | | 286+ | - | 6.7% | 40.0% | 7.6% | 63.2% |
| | | | | | 271+ | - | 10.0% | 50.0% | 5.5% | 68.7% |

| | | | | | | | | | | |
|---|---|---|---|---|---|---|---|---|---|---|
| Self-derived (men & women) | Dev | Balzer (2020) | SEARCH (Risk group model) | - | - | - | 20.0% | 20.0% | 8.0% | 8.0% |
| | | | | | - | - | 10.0% | 30.0% | 0.0% | 8.0% |
| | | | | | - | - | 10.0% | 40.0% | 0.0% | 8.0% |
| | | | | | - | - | 5.0% | 45.0% | 50.0% | 58.0% |
| | Dev | | SEARCH (Logistic model) | - | - | - | 20.0% | 20.0% | 40.0% | 40.0% |
| | | | | | - | - | 10.0% | 30.0% | 15.0% | 55.0% |
| | | | | | - | - | 10.0% | 40.0% | 13.0% | 68.0% |
| | | | | | - | - | 5.0% | 45.0% | 0.0% | 68.0% |
| | Dev | | SEARCH (Machine-learning) | - | - | - | 20.0% | 20.0% | 52.0% | 52.0% |
| | | | | | - | - | 10.0% | 30.0% | 13.0% | 65.0% |
| | | | | | - | - | 10.0% | 40.0% | 9.0% | 74.0% |
| | | | | | - | - | 5.0% | 45.0% | 4.0% | 78.0% |
| Self-derived (men) | Dev | Roberts ¶ (2021) | ACDIS (women) | - | 5th quintile | 2.7 | 20.0% | 20.0% | - | - |
| | | | | | 4th quintile | 1.6 | 20.0% | 40.0% | - | - |
| | | | | | 3rd quintile | 0.6 | 20.0% | 60.0% | - | - |
| | | | | | 2nd quintile | 0.5 | 20.0% | 80.0% | - | - |
| | | | | | 1st quintile | 0.3 | 20.0% | 100.0% | - | - |
| Self-derived (men) | Dev | Roberts ¶ (2021) | ACDIS (women) | - | 5th quintile | 6.5 | 20.0% | 20.0% | - | - |
| | | | | | 4th quintile | 3.8 | 20.0% | 40.0% | - | - |
| | | | | | 3rd quintile | 2.9 | 20.0% | 60.0% | - | - |
| | | | | | 2nd quintile | 1.9 | 20.0% | 80.0% | - | - |
| | | | | | 1st quintile | 0.7 | 20.0% | 100.0% | - | - |

¶ Incidence are presented graphically in Burgess (2018) [11], Rosenberg (2020) [13], Peebles (2020) [9], and Roberts (2021) [19].

* Self-reported STIs histories or symptoms were used as proxies for curable STIs in Burgess (2017) [27] and Rosenberg (2020) [13] and for curable STIs in Burgess (2018) [11].

**Figure S1.** **Risk of bias assessment (A) and concerns for applicability (B) for the model development (i) and validation (ii) studies.**

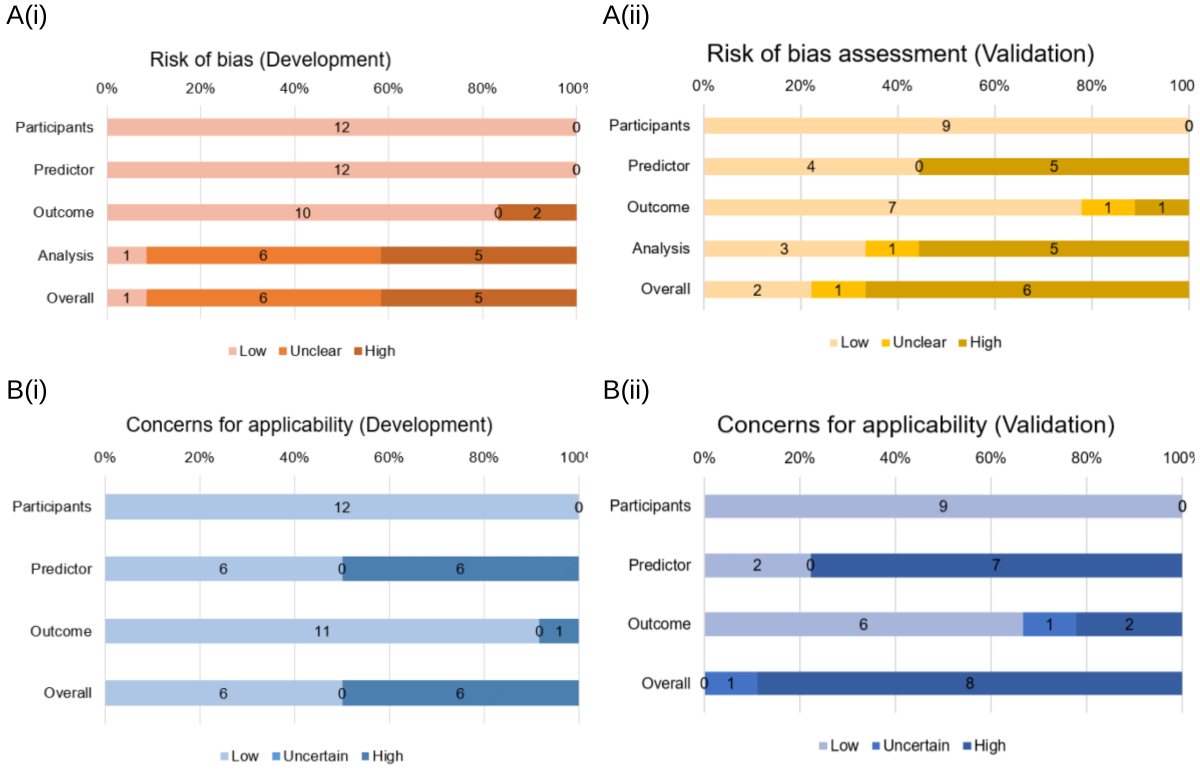

**Appendix III. PRIMSA 2020 Abstract Checklist**

| Topic | No. | Item | Reported? |
|---|---|---|---|
| **TITLE** | | | |
| **Title** | 1 | Identify the report as a systematic review. | Yes |
| **BACKGROUND** | | | |
| **Objectives** | 2 | Provide an explicit statement of the main objective(s) or question(s) the review addresses. | Yes |
| **METHODS** | | | |
| **Eligibility criteria** | 3 | Specify the inclusion and exclusion criteria for the review. | Yes |
| **Information sources** | 4 | Specify the information sources (e.g. databases, registers) used to identify studies and the date when each was last searched. | No |
| **Risk of bias** | 5 | Specify the methods used to assess risk of bias in the included studies. | Yes |
| **Synthesis of results** | 6 | Specify the methods used to present and synthesize results. | Yes |
| **RESULTS** | | | |
| **Included studies** | 7 | Give the total number of included studies and participants and summarise relevant characteristics of studies. | Yes |
| **Synthesis of results** | 8 | Present results for main outcomes, preferably indicating the number of included studies and participants for each. If meta-analysis was done, report the summary estimate and confidence/credible interval. If comparing groups, indicate the direction of the effect (i.e. which group is favoured). | Yes |
| **DISCUSSION** | | | |
| **Limitations of evidence** | 9 | Provide a brief summary of the limitations of the evidence included in the review (e.g. study risk of bias, inconsistency and imprecision). | No |
| **Interpretation** | 10 | Provide a general interpretation of the results and important implications. | Yes |
| **OTHER** | | | |
| **Funding** | 11 | Specify the primary source of funding for the review. | Yes |
| **Registration** | 12 | Provide the register name and registration number. | Yes |

*From:* Page MJ, McKenzie JE, Bossuyt PM, Boutron I, Hoffmann TC, Mulrow CD, et al. The PRISMA 2020 statement: an updated guideline for reporting systematic reviews. MetaArXiv. 2020, September 14. DOI: 10.31222/osf.io/v7gm2. For more information, visit: www.prisma-statement.org

#### Appendix IV. PRISMA 2020 Main Checklist

| Topic | No. | Item | Location where item is reported |
|---|---|---|---|
| **TITLE** | | | |
| **Title** | 1 | Identify the report as a systematic review. | p.1 |
| **ABSTRACT** | | | |
| **Abstract** | 2 | See the PRISMA 2020 for Abstracts checklist | |
| **INTRODUCTION** | | | |
| **Rationale** | 3 | Describe the rationale for the review in the context of existing knowledge. | p.6-7 |
| **Objectives** | 4 | Provide an explicit statement of the objective(s) or question(s) the review addresses. | p.7 |
| **METHODS** | | | |
| **Eligibility criteria** | 5 | Specify the inclusion and exclusion criteria for the review and how studies were grouped for the syntheses. | p.8 |
| **Information sources** | 6 | Specify all databases, registers, websites, organisations, reference lists and other sources searched or consulted to identify studies. Specify the date when each source was last searched or consulted. | p.8 |
| **Search strategy** | 7 | Present the full search strategies for all databases, registers and websites, including any filters and limits used. | p.8; Appendix I |
| **Selection process** | 8 | Specify the methods used to decide whether a study met the inclusion criteria of the review, including how many reviewers screened each record and each report retrieved, whether they worked independently, and if applicable, details of automation tools used in the process. | p.8 |
| **Data collection process** | 9 | Specify the methods used to collect data from reports, including how many reviewers collected data from each report, whether they worked independently, any processes for obtaining or confirming data from study investigators, and if applicable, details of automation tools used in the process. | p.8 |
| **Data items** | 10a | List and define all outcomes for which data were sought. Specify whether all results that were compatible with each outcome domain in each study were sought (e.g. for all measures, time points, analyses), and if not, the methods used to decide which results to collect. | p.8; Appendix II |
| | 10b | List and define all other variables for which data were sought (e.g. participant and intervention characteristics, funding sources). Describe any assumptions made about any missing or unclear information. | p.8; Appendix II |
| **Study risk of bias assessment** | 11 | Specify the methods used to assess risk of bias in the included studies, including details of the tool(s) used, how many reviewers assessed each study and whether they worked independently, and if applicable, details of automation tools used in the process. | p.8-9 |
| **Effect measures** | 12 | Specify for each outcome the effect measure(s) (e.g. risk ratio, mean difference) used in the synthesis or presentation of results. | p.9 |

| Topic | No. | Item | Location where item is reported |
|---|---|---|---|
| **Synthesis methods** | 13a | Describe the processes used to decide which studies were eligible for each synthesis (e.g. tabulating the study intervention characteristics and comparing against the planned groups for each synthesis (item 5)). | p.9 |
| | 13b | Describe any methods required to prepare the data for presentation or synthesis, such as handling of missing summary statistics, or data conversions. | p.9 |
| | 13c | Describe any methods used to tabulate or visually display results of individual studies and syntheses. | p.9 |
| | 13d | Describe any methods used to synthesize results and provide a rationale for the choice(s). If meta-analysis was performed, describe the model(s), method(s) to identify the presence and extent of statistical heterogeneity, and software package(s) used. | p.9 |
| | 13e | Describe any methods used to explore possible causes of heterogeneity among study results (e.g. subgroup analysis, meta-regression). | N/A |
| | 13f | Describe any sensitivity analyses conducted to assess robustness of the synthesized results. | N/A |
| **Reporting bias assessment** | 14 | Describe any methods used to assess risk of bias due to missing results in a synthesis (arising from reporting biases). | N/A |
| **Certainty assessment** | 15 | Describe any methods used to assess certainty (or confidence) in the body of evidence for an outcome. | N/A |
| **RESULTS** | | | |
| **Study selection** | 16a | Describe the results of the search and selection process, from the number of records identified in the search to the number of studies included in the review, ideally using a flow diagram. | p.10 |
| | 16b | Cite studies that might appear to meet the inclusion criteria, but which were excluded, and explain why they were excluded. | Figure 1 |
| **Study characteristics** | 17 | Cite each included study and present its characteristics. | p.10 |
| **Risk of bias in studies** | 18 | Present assessments of risk of bias for each included study. | Table S4 |
| **Results of individual studies** | 19 | For all outcomes, present, for each study: (a) summary statistics for each group (where appropriate) and (b) an effect estimate and its precision (e.g. confidence/credible interval), ideally using structured tables or plots. | Figure 2-4; Table 2-3 |
| **Results of syntheses** | 20a | For each synthesis, briefly summarise the characteristics and risk of bias among contributing studies. | Figure 2-4 |
| | 20b | Present results of all statistical syntheses conducted. If meta-analysis was done, present for each the summary estimate and its precision (e.g. confidence/credible interval) and measures of statistical heterogeneity. If comparing groups, describe the direction of the effect. | Figure 2-4 |
| | 20c | Present results of all investigations of possible causes of heterogeneity among study results. | p.12-13 |
| | 20d | Present results of all sensitivity analyses conducted to assess the robustness of the synthesized results. | N/A |

| Topic | No. | Item | Location where item is reported |
|---|---|---|---|
| **Reporting biases** | 21 | Present assessments of risk of bias due to missing results (arising from reporting biases) for each synthesis assessed. | N/A |
| **Certainty of evidence** | 22 | Present assessments of certainty (or confidence) in the body of evidence for each outcome assessed. | N/A |
| **DISCUSSION** | | | |
| **Discussion** | 23a | Provide a general interpretation of the results in the context of other evidence. | p.14-16 |
| | 23b | Discuss any limitations of the evidence included in the review. | p.15-16 |
| | 23c | Discuss any limitations of the review processes used. | p.16 |
| | 23d | Discuss implications of the results for practice, policy, and future research. | p.16 |
| **OTHER INFORMATION** | | | |
| **Registration and protocol** | 24a | Provide registration information for the review, including register name and registration number, or state that the review was not registered. | p.2, 10 |
| | 24b | Indicate where the review protocol can be accessed, or state that a protocol was not prepared. | p.2, 10 |
| | 24c | Describe and explain any amendments to information provided at registration or in the protocol. | N/A |
| **Support** | 25 | Describe sources of financial or non-financial support for the review, and the role of the funders or sponsors in the review. | p. 28 |
| **Competing interests** | 26 | Declare any competing interests of review authors. | p. 28 |
| **Availability of data, code and other materials** | 27 | Report which of the following are publicly available and where they can be found: template data collection forms; data extracted from included studies; data used for all analyses; analytic code; any other materials used in the review. | N/A |

*From:* Page MJ, McKenzie JE, Bossuyt PM, Boutron I, Hoffmann TC, Mulrow CD, et al. The PRISMA 2020 statement: an updated guideline for reporting systematic reviews. MetaArXiv. 2020, September 14. DOI: 10.31222/osf.io/v7gm2. For more information, visit: [www.prisma-statement.org](http://www.prisma-statement.org)
